# Single-Case Design for Network-based BCI Rehabilitation: Individual Learning Trajectories and Effects of Motor Imagery Timing in Stroke Patients

**DOI:** 10.64898/2026.10.21.26366015

**Authors:** Camile Bousfiha, Camille Gabot, Diana Baili, Erin Soller, Marion Couton, Grace Adwane, Michaёl Obadia, Chiara Zavanone, Sophie Dupont, Eléonore Bayen, Laurent Bougrain, Paolo Bartolomeo, Fabrizio de Vico Fallani

## Abstract

**Background:** Motor imagery (MI) brain-computer interfaces (BCIs) are increasingly investigated for promoting motor recovery after stroke. However, it remains unclear how BCI control improves through repeated training in stroke patients and whether the timing of MI decoding systematically influences this trajectory.

**Methods:** To address this question, we implemented a single-case experimental design involving five chronic stroke patients enrolling in a longitudinal electroen-cephalography MI-BCI training involving their affected hand. Across 18 sessions conducted over six weeks, the onset of a fixed-duration MI decoding window was randomized to occur 1, 2, or 3 s after cue onset according to an alternating-treatments single-case design. Notably, the BCI combined spectral power and functional connectivity features to classify MI states.

**Results:** The MI classification timing had no significant effect on BCI accuracy, fatigue, or spasticity. BCI accuracy increased across training in four of five patients and was accompanied by patient-specific longitudinal changes in local spectral activity and functional connectivity. These convergent trajectories were consistent with acquisition of BCI control, but were not accompanied by clinically meaningful improvement in motor function.

**Conclusions:** Varying the onset of the MI decoding window provided no systematic benefit for BCI control. Instead, differences in neurophysiological trajectories across patients indicate that MI decoding parameters need to be personalized and reassessed throughout the training. Overall, these findings inform more individualized and effective BCI settings for neuromotor stroke rehabilitation. **Trial registration:** ClinicalTrials.gov, NCT06951035, registered 30 April 2025

## 1 Introduction

Motor imagery (MI) brain-computer interfaces (BCIs) are widely studied for upper-limb rehabilitation after stroke and are of particular interest for patients whose voluntary motor function is too limited to engage in conventional active practice [1]. Nevertheless, numerous experimental paradigms exist, making it difficult to disentan-gle the contribution of individual parameters and to conclude on best practices [2]. Consequently, protocol parameters are often fixed across participants even though the behavioral and neurophysiological characteristics relevant to BCI control may differ substantially between patients. This issue is particularly relevant after stroke. Lesion location and extent, severity of the motor deficit, cognitive comorbidity and stage of the recovery trajectory differ from one patient to another, and influence the individual response to a given BCI treatment.

These patient-specific factors can shape the neurophysiological expression of motor imagery, including the oscillatory frequency band in which the discriminant response emerges [3, 4] and how this response evolves over training. As a result, electrophysiology event-related responses within a single cohort can even go in opposite directions [5], so that a group-average effect may not describe the trajectory of an individual patient [6]. As a matter of fact, designing a BCI paradigm for stroke rehabilitation often implies choices inherited from local practices, technical and algorithmic constraints, as well as accommodations to patients’ needs [7]. Frameworks for instruction and user training do exist for healthy volunteers, but no equivalent guidance covers rehabilitation after stroke [8, 9].

An important aspect of rehabilitation is the time spent practicing and repeating targeted motor tasks. Conventional therapies deliver around 32 repetitions per session/day, whereas animal models suggest around 500 repetitions are needed to induce cortical synaptic changes [10]. Accordingly, BCI protocols should be intensive in terms of number of MI repetitions/trials and number of consecutive sessions. At the same time, any BCI protocol should also avoid excessive fatigue and drowsiness in patients, knowing that the EEG setup already takes about 20% of the entire session [11]. Another crucial factor is the duration of the trial associated with the MI task, typically a trade-off between 3 [12] and 10 seconds or more [13]. Longer durations may account for changes in MI responses associated with motor system alterations after stroke [14, 15]. Shorter durations are mainly intended to shorten the overall session, made of several consecutive trials, and therefore diminish patients’ fatigue.

Finally, most of the approaches in the BCI field have focused on univariate local features, such as EEG power spectra and even-related dynamics changes. However, stroke is a network-level disorder where both MI and recovery depends on the reorganization of intra- and interhemispheric interactions involving primary and associative brain areas [16–18]. Approaches based on functional connectivity [19–21] and covariance[22] between EEG signals are therefore more grounded candidates to provide richer network-based feedback in MI-BCI after stroke.

Motivated by the aforementioned considerations, we realized a functional electrical stimulation (FES)-controlled EEG network-based MI-BCI study involving 5 chronic stroke patients over 18 equally-spaced sessions spanning 6 weeks of training. Notably, we manipulated the time allocated for the MI task across sessions according to a single-case experimental design (SCED) with randomized alternating treatment, so that patients served as their own control [23]. We evaluated whether this intense protocol can be tolerated by patients, whether their online BCI accuracy improves over the training, in association with specific brain activity and network changes, and whether the MI timing had a significant overall impact.

## 2 Methods

### 2.1 Experimental protocol

Five chronic BCI-naive stroke patients were included in the study. Inclusion criteria were chronic stroke (*≥* 6 months after a first-ever lesion), residual distal upper-limb impairment (UE-FMA *<* 53) [24], and visible active finger flexion (MRC *≥* 2) [25]. Exclusion criteria included insufficient command comprehension (BDAE *<* 12) [26], fixed finger or wrist flexor spasticity (mAS = 4) [27], botulinum toxin injection in the forearm or hand within the previous 12 weeks, and any contraindication to FES or EEG recording Other eligibility criteria were reported in the **Supplementary Methods (Methods S1)**, together with the full inclusion assessment battery tests (**Methods S2.1**). No screening for BCI aptitude was applied

This study was part of a larger clinical trial (NCT06951035) that was approved by an independent French research ethics committee (CPP Sud-Mediterrańee I, ID-RCB 2024-A00230-47) and authorised by the French National Agency for Medicines and Health Products Safety. All patients gave written informed consent in accordance with the Declaration of Helsinki.

#### 2.1.1 FES-controlled MI-BCI training

Upon enrollment, patients underwent 18 BCI training sessions over six weeks (spaced 1-2 days apart), followed by a three-month post-intervention assessment. Before each session, participants rated their mood, stress and sleep quality on a tactile visual support with a continuous scale from 0 to 100. Fatigue was rated on the same scale before, during and after each session. Spasticity was measured with the mAS scale on the thumb, the index, the other fingers and the wrist, before and after each session. Each segment was graded on the six-level modified Ashworth scale [27], with grade 1+ scored as 2 and higher grades shifted accordingly, and the four segment scores were summed into a global score ranging from 0 to 20. Upper-limb motor functions were measured with the FMA scale before the first and last session. All behavioral and clinical scales were retaken a final time at the follow-up visit. Each session was conducted in a quiet shielded controlled environment. Patients were comfortably seated in front of a screen placed horizontally on a height-adjustable table, with the paretic upper limb supported on a mobile armrest to ensure optimal positioning and minimize fatigue (**Fig. 1**). FES was delivered through the FESIA Grasp system [28], positioned on the paretic forearm and calibrated before training to produce a standardized and functional hand opening, more details about FESIA calibration are provided in **Methods S2.2**.

**Fig. 1.**
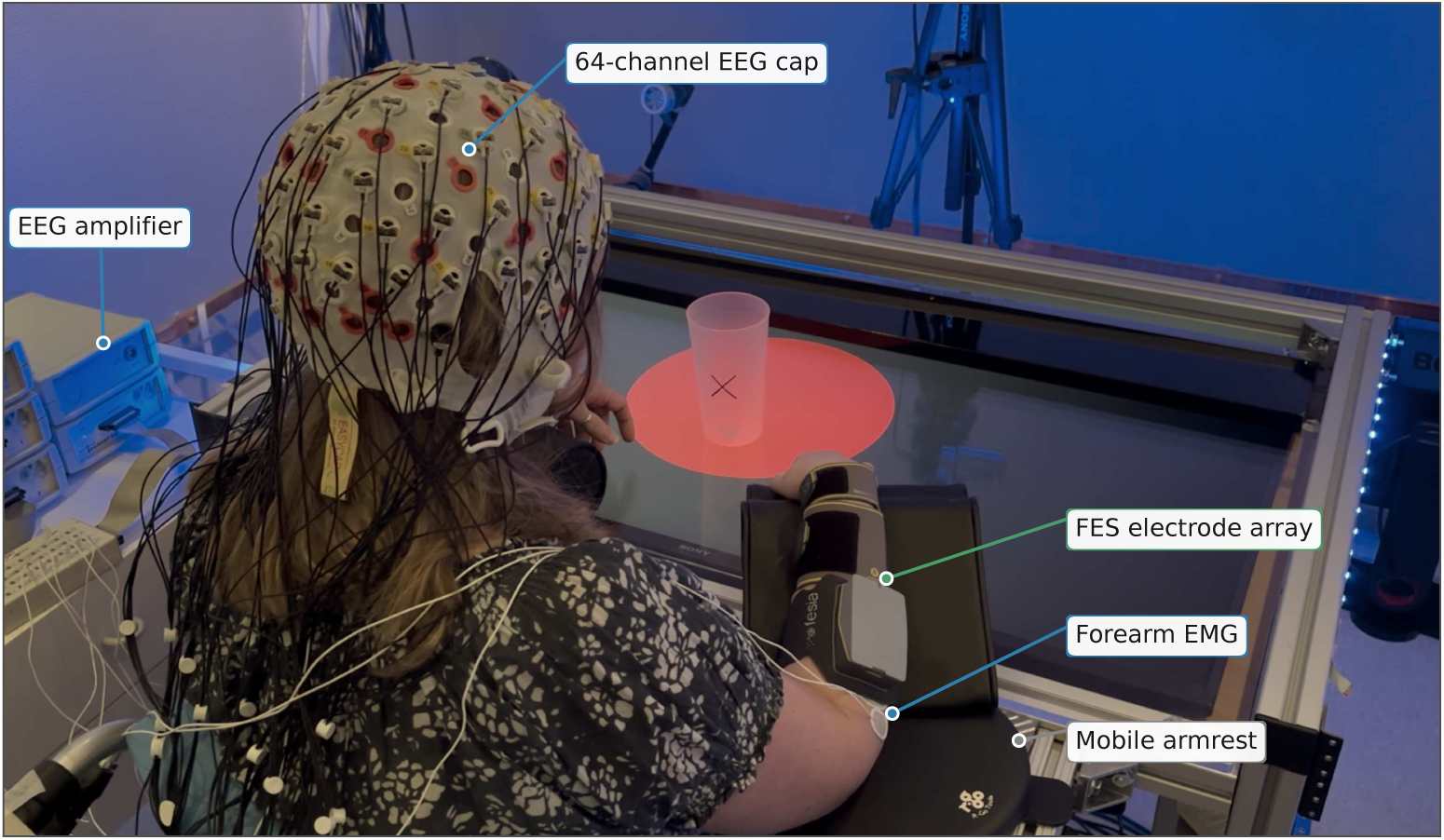
Experimental setup. BCI training sessions were conducted in a quiet, shielded room under dim ambient lighting. The patient sat in front of a horizontally positioned screen mounted on a height-adjustable table, which displayed the entire task interface. During rest phases, a grey empty circle was presented; during the cue period, the ring was filled in red, as illustrated here. A target cup was positioned on the screen in front of the patient so that imagined hand opening was directed toward a concrete object. EEG signals were recorded using a 64-channel cap connected to an amplifier. The paretic forearm rested on a mobile arm support and was fitted with the FESIA Grasp electrode array, together with surface EMG electrodes used to verify the absence of voluntary muscle contraction during the motor imagery phases.

Patients were instructed to imagine opening their paretic hand in a sustained manner. Specifically, patients were invited to focus on the internal sensation of movement, including muscle contraction and joint displacement (kinesthetic MI), and to visualize finger extension from a first-person perspective (visual MI). A target cup was placed on the table in front of the patients to help them perform the task in an ecological and functional way. Standardized instructions were given before each session and included a brief breathing exercise followed by guided MI. A detailed script was used during the first three sessions and an abbreviated version thereafter (**Methods S3.1**). Each session consisted of nine consecutive runs of 18 MI trials, corresponding to 162 MI trials in total. The first run was used for *calibration* and always triggered FES, whereas the subsequent eight *testing* comprised 144 closed-loop trials in which FES depended on BCI classification (**Fig. 2**). Each trial began with a 5 s Rest (baseline) phase cued by an open circle on the table-screen, followed by a MI phase, triggered by a red circle, that lasted between 4, 5 or 6 s.

**Fig. 2.**
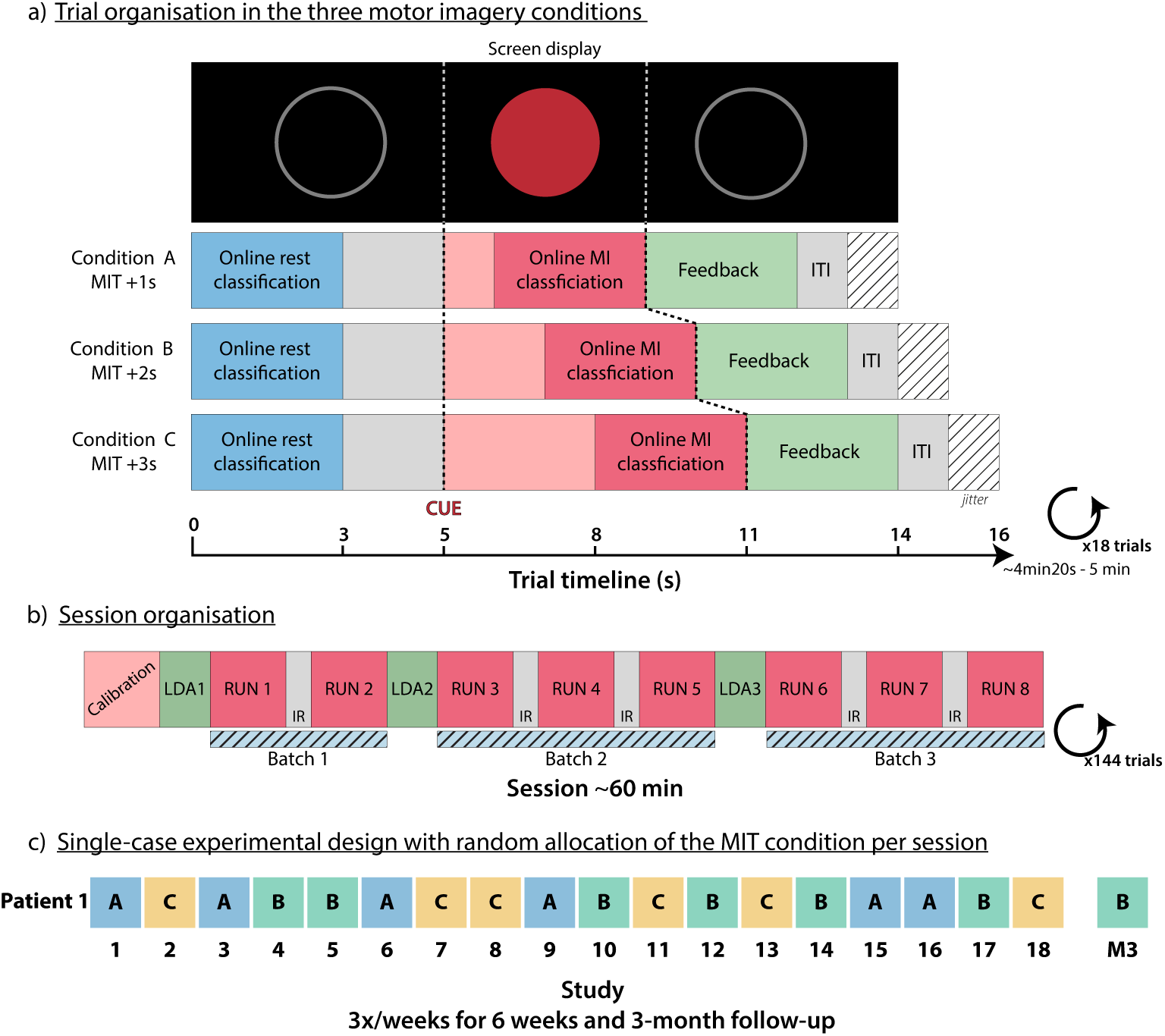
Study design. **a)** The screen showed a grey circle, which filled red throughout the cue window. The 3-s online rest classification started at the beginning of the trial and ended 2 s before the cue, at which patients were instructed to perform sustained hand-opening imagery. The 3-s online MI classification (MIT) started 1, 2, or 3 s after the cue. FES feedback was delivered for 3 sec at the end of the cue window if MI decoding succeeded. A jitter of 1 or 2 s served as the inter-trial interval (ITI) to limit anticipation of the next cue. **b)** Each session comprised three batches of BCI training. Three decoders (LDA1 to LDA3) were trained in turn, each on the runs preceding the batch it controlled. During each run, the experimenter monitored the decoding of each class in real time, together with EEG artifacts and the EMG signal. Between runs, an inter-run break (IR) was taken at the patient’s or the experimenter’s discretion. Between batches, feature evolution was analyzed, and features were selected and trained semi-automatically with the HappyFeat software, in about 3 min. During this time, the experimenter debriefed with the patient, based on their own report, the real-time classification and the feature drift. **c)** Sessions were held three times a week, at the same time of day, for six weeks, with a 3-month follow-up. The allocation of the MIT condition was randomized *a priori* and balanced across the three conditions, six sessions each, for every patient.

In the final 3-second feedback phase, the FES was delivered only when the MI was correctly detected by the classifier. Random inter-trial intervals between 1 and 2 s were introduced to reduce temporal predictability and anticipation. This trial structure was intended to allow smooth transitions between phases and avoid overly long trials that could increase fatigue or reduce attentional engagement. At the end of each run, the experimenter debriefed with the patient through a discussion, detailed in **Methods S3.2**.

Online BCI performance was measured in terms accuracy, defined as the number of correctly classified Rest and MI phases over two times the total number of trials.

#### 2.1.2 Single-case experimental design

To evaluate whether the timing of MI decoding affected BCI control, we implemented a randomized alternating-treatments single-case experimental design [23] (Fig. 2b). Three MI classification timing (MIT) conditions were defined by shifting the onset of a fixed 3-s decoding window to 1 (condition A), 2 (condition B) or 3 (condition C) seconds after cue onset. Accordingly, the entire cue period lasted 4, 5, or 6 s so that classification always used the final 3 s of the MI phase. These conditions were assigned using a balanced randomization scheme, ensuring equal presentation across the 18 sessions (**Fig. 2**). Patients, but not experimenters, were blinded to the MI duration tested in each session.

### 2.2 Online EEG recording and processing

#### 2.2.1 EEG acquisition

EEG was recorded with a 64-channel BrainAmp system (Brain Products, Germany) with active electrodes and a 500 Hz sampling frequency, following a 10-20 montage and referenced to the mastoids (**Supplementary Figures (Fig. S1)**). Electrode impedance was lowered and maintained below 10 kΩ at the beginning of each session. Bilateral forearm electromyography was recorded and monitored during the session to ensure that muscular contractions were not present during the MI phases. The following signal processing and statistical analysis were carried out with the MNE-Python library, while online feature extraction and decoder training used the HappyFeat software [29]. The online BCI pipeline and its interaction with the FES device was implemented using the OpenViBE software [30]. A notch filter at 50 Hz and its harmonics with a common average reference was applied on the signal. The frontotemporal electrodes FT9 and FT10, as well as the inion electrode Iz, were excluded from the analyses.

#### 2.2.2 Feature extraction

To ensure comparability, all the EEG features used to control the BCI were calculated on the basis of a fixed 3-second window. During the Rest phase, this window corresponded to the first three seconds; during the MI phase, the position of the window depended on the SCED, i.e. 1 s after the cue for 4 s MI duration, 2 s after the cue for 5 s MI duration, and 3 s after the cue for 6 s MI duration (**Fig. 2b**).

On the segmented windows, we estimated the power spectral density (*ϕ*) over 0.25 s epochs shifted by 0.161 s. To get the most reliable estimate for such short-time windows, we used an autoregressive Burg model of order 24 with a spectral resolution of 2 Hz. The resulting estimates were eventually averaged to get one robust measure for the 3-second window.

Functional connectivity was calculated on sliding 0.5 s epochs shifted by 0.32 s to cover the entire 3-second window. Specifically, we computed the imaginary coherence [31] to measure phase-synchronized interactions between brain signals while suppressing spurious zero-lag correlations due to volume conduction. This measure has been widely used in MI-BCI studies[17, 19, 20, 32]. The resulting brain connectivity network can be represented as a full weighted matrix

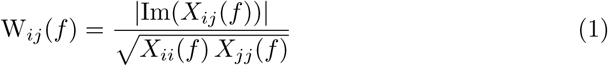

where *X_ij_* is the cross-spectrum between the EEG signals recorded at electrodes *i* and *j* at frequency *f* and *X_ii_*, *X_jj_* are the corresponding autospectra. Similarly to power spectra, we used a bivariate autoregressive Burg model of order 24 with a spectral resolution of 2 Hz to estimate the auto- and cross-spectra. The resulting connectivity matrices *W* were eventually averaged to get one robust network for the 3-second window.

Using the network neuroscience formalism, we calculated the node strength (*s*) as the sum of the weighted connections (or links) incident to a given EEG electrode *i* (i.e. a node of the network)

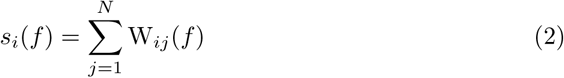

where *N* is the total number of nodes. By locally integrating information from the other nodes, node strengths make it possible to identify the most strongly connected electrodes in the EEG network, thus reducing complexity and favoring interpretability [19, 20].

#### 2.2.3 Feature selection, classifier, and FES control

We adopted a semi-automatic procedure to select the most relevant brain activity (power spectrum, *ϕ*) and network (node strength connectivity, *s*) features.

Candidate features were first visualized on a *r*^2^ channel-frequency map [33] computed between MI and Rest phases, for *ϕ* and *s* features separately. They were then ranked according to their magnitude, only considering the sensorimotor electrodes (FC, C, CP) (**Fig. S1**) within the alpha (8 *−* 12 Hz) and beta range (13 *−* 30 Hz). A regularized linear discriminant analysis (LDA) classifier [34] was separately trained on three different balanced combinations of power and network features, using the best one, two or three *ϕ*-*s* pairs. The combination achieving the highest classification accuracy was selected. This feature selection process was repeated at three different time points during the session to allow the classifier to adapt to possible non-stationarities arising from mental fatigue, mental strategy changes and signal drift (**Fig. 2b**). Specifically, the first classifier was trained on the initial *calibration* run and used for testing runs 1 *−* 2. It was then retrained using data from testing runs 1 *−* 2 and applied to testing runs 3 *−* 5. Finally, the classifier was retrained on data from testing runs 3 *−* 5 and applied to *testing* runs 6 *−* 8.

### 2.3 Offline performance and neurophysiological measures

#### 2.3.1 Offline BCI performance

Three offline metrics of MI-BCI skill were computed for each run [35]. Run-wise cross-validated accuracy was computed using four-fold cross-validation within each run, with common spatial patterns and a linear discriminant analysis over 8 *−* 30 Hz, ensuring a constant amount of training data throughout the protocol.

To complement this classifier-dependent measure, we quantified two classifier-free properties of the covariance distributions: their separation and their within-class consistency. Rest distinctiveness was the Riemannian distance between the imagery and rest covariance means divided by their mean dispersion, thereby measuring how distinctly MI-related activity departed from rest. Class stability was defined as the inverse dispersion of the imagery class around its own mean, capturing the reproducibility of MI-related activity across trials. Covariances were estimated on the unreferenced raw signal.

#### 2.3.2 Event-related spectral dynamics

Relative power change was quantified separately for MI and Rest at each frequency bin as (*X −R*)*/R×* 100, where *X* denotes the EEG power computed over the same 3-s MI or Rest analysis window used for online classification. The reference *R* was a 1-s window, placed just after the rest period and 1 s before the cue to keep anticipation out of it [36]. Thus, negative values denote an event-related desynchronization (ERD), positive values an event-related synchronization (ERS).

First, to characterize the temporal dynamics of the ERD and ERS responses and their evolution across training, we identified the most discriminant ERD and ERS site in each session from the signed *r*^2^ maps. The *r*^2^ map of each session was tested by threshold-free cluster enhancement (TFCE) (*E* = 2*/*3, *H* = 2, 2000 sign flips) [37, 38] (**Fig. S2.A**). For each session, the significant channel-frequency bin with the largest negative value was taken as the ERD site, and the bin with the largest positive value as the ERS site; where the map held no significant bin of a given sign, the corresponding site was left undefined. Significance over time was then established at each site using the studentized bootstrap of Graimann et al. [39] (1000 resamples per window, *α* = 0.05), pooling the trials of the session. The analysis used overlapping bands whose width grew with frequency (2, 4 then 8 Hz), with the lower cutoff stepping by 1 Hz from 8 to 31 Hz (**Fig. S2.B**). To guard against noise, isolated significant windows were discarded.

Four metrics were then derived from each session’s response. Amplitude was the area under the relative power change curve. Inter-trial variability was the standard deviation of that per-trial amplitude across the trials of the session. Duration was the fraction of significant windows, and latency, the time of the earliest of them, left undefined when the analysis period began with a significant window already present, as this would reflect anticipation rather than a genuine response to the cue. Each metric was computed over a specific window: amplitude over the 3-s rest and imagery windows; inter-trial variability over the imagery window; duration over the 3-s rest window and on the cued window (condition duration plus the 3-s imagery window); and latency over the cued window (**Fig. S2.C**).

Second, to characterize longitudinal changes in MI-Rest discriminability, session slopes were estimated from the signed *r*^2^ maps of spectral power *ϕ* and node strength *s* over the alpha and beta bands. The analysis was restricted to sensorimotor electrodes. Connected components were formed separately among channel-frequency bins of each sign, so that each cluster was homogeneous in direction; negative clusters corresponded to MI*<*Rest responses and positive clusters to MI*>*Rest responses.

#### 2.3.3 Brain network properties

To assess how MI altered the brain network at the single connection level, we calculated the signed coefficient of determination *r*^2^ between the EEG network link weights in MI and Rest windows, such that negative values denote MI decrements.

To measure the contribution of within-hemisphere segregation and between-hemisphere integration, we averaged these values for the affected hemisphere (*D*_AH_), the unaffected hemisphere (*D*_UH_), and between hemispheres [17, 18]. The hemispheric balance was assessed by the laterality index [17, 19] :

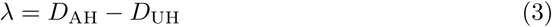

positive values indicating a stronger contrast on the lesioned side (toward coupling, or away from decoupling).

To characterize the topological specificity of these changes relative to the motor network, edges were further divided into three classes: inner motor edges, connecting two motor electrodes; outer motor edges, connecting a motor electrode to a non-motor electrode; and extramotor edges, connecting two non-motor electrodes as represented in **Fig. S3**. Longitudinal changes in signed *r*^2^ were then estimated separately for each edge class. A negative slope denotes progressive MI decoupling relative to Rest.

### 2.4 Statistical analysis

The primary outcome was the longitudinal evolution of online BCI accuracy. Complementary outcomes included offline BCI performance, ERD/ERS dynamics, longitudinal *r*^2^ clusters of power spectral density (*ϕ*) and node strength (*s*), network intensities, fatigue, and spasticity.

Session-level outcomes were analyzed using an analysis of covariance (ANCOVA) including three terms: session number, MIT, and fatigue reported on arrival, with patient-specific intercepts in the group-level analyses. Session number represented the longitudinal trajectory across training. MIT was entered as a linear term corresponding to decoding-window onsets of 1, 2, or 3 s after cue onset. Fatigue was included as a nuisance covariate when it varied across sessions. The two effects of interest had different inferential status. MIT was experimentally randomized and was therefore tested by permuting the MIT assignments according to the randomization scheme implemented in the study, following the Draper-Stoneman principle [40], thereby pre-serving the design-based randomization inference. Session number was observational rather than randomized and was tested conditionally on the remaining model using Freedman-Lane permutation of the residuals from the reduced model [41, 42].

Inference was based on 2000 permutations of a heteroscedasticity-robust Wald statistic [43]. Permutations were restricted within each patient, so that observations were only exchanged among sessions from the same patient [44]. The observed configuration was included in the permutation distribution, yielding a minimum attainable *p*-value of 1*/*2001 *≈* 0.0005. Each 95% confidence interval was obtained by inversion of the corresponding permutation test.

Offline BCI metrics were analyzed as complementary outcomes to determine whether longitudinal changes persisted when performance was evaluated independently of online decoder adaptation. Tolerance was assessed from fatigue and spasticity, testing both their longitudinal evolution and their within-session change. ERD/ERS temporal dynamics were analyzed using six session-level outcomes: MI amplitude, Rest amplitude, MI inter-trial variability, MI duration, Rest duration, and MI latency.

Neurophysiological changes were examined at two complementary levels. First, longitudinal changes in the signed *r*^2^ maps of spectral power *ϕ* and node strength *s* characterized the local and node-level evolution of MI-Rest discriminability. Second, edge-level functional connectivity was used to characterize longitudinal changes in motor-network organization. MI and Rest connectivity were subsequently modeled separately to determine whether longitudinal changes in the MI-Rest contrast arose from changes during MI, Rest, or both.

Multiple-comparison correction depended on the outcome family. For the six temporal ERD/ERS outcomes, the session and MIT effects were corrected separately using the Benjamini-Hochberg procedure (*p*_BH_) [45], within each patient and at the group level. For *r*^2^ maps of spectral power and node strength, the session and MIT effects were tested separately, with family-wise error controlled at 5% using permutation-based threshold-free cluster enhancement. For edge-level network measures, the session and MIT effects were corrected separately using the Benjamini-Hochberg procedure across the nine combinations of network subdivision (inner motor, outer motor, and extramotor) and hemispheric contribution (affected hemisphere, unaffected hemisphere, and interhemispheric), and across the three corresponding laterality indices.

## 3 Results

### 3.1 Behavioral outcome and BCI performance

Five patients were included with no drop-outs. Their characteristics are given in **Table 1**. All patients were chronic, between 12 and 63 months after their first ever stroke. Three patients had residual fingers flexion without voluntary fingers extension (P1,P3,P4). One patient (P1) presented a non-fluent aphasia classified as global, its severity carried by expression, comprehension being sufficiently preserved to meet the inclusion threshold on command execution. Two protocol deviations occurred: session 18 of P1 delivered 6 s of MI instead of the 5 s of its assigned condition, and session 2 of P3 delivered 4 s instead of 5 s. Both were analyzed under the duration actually delivered.

**Table 1.**
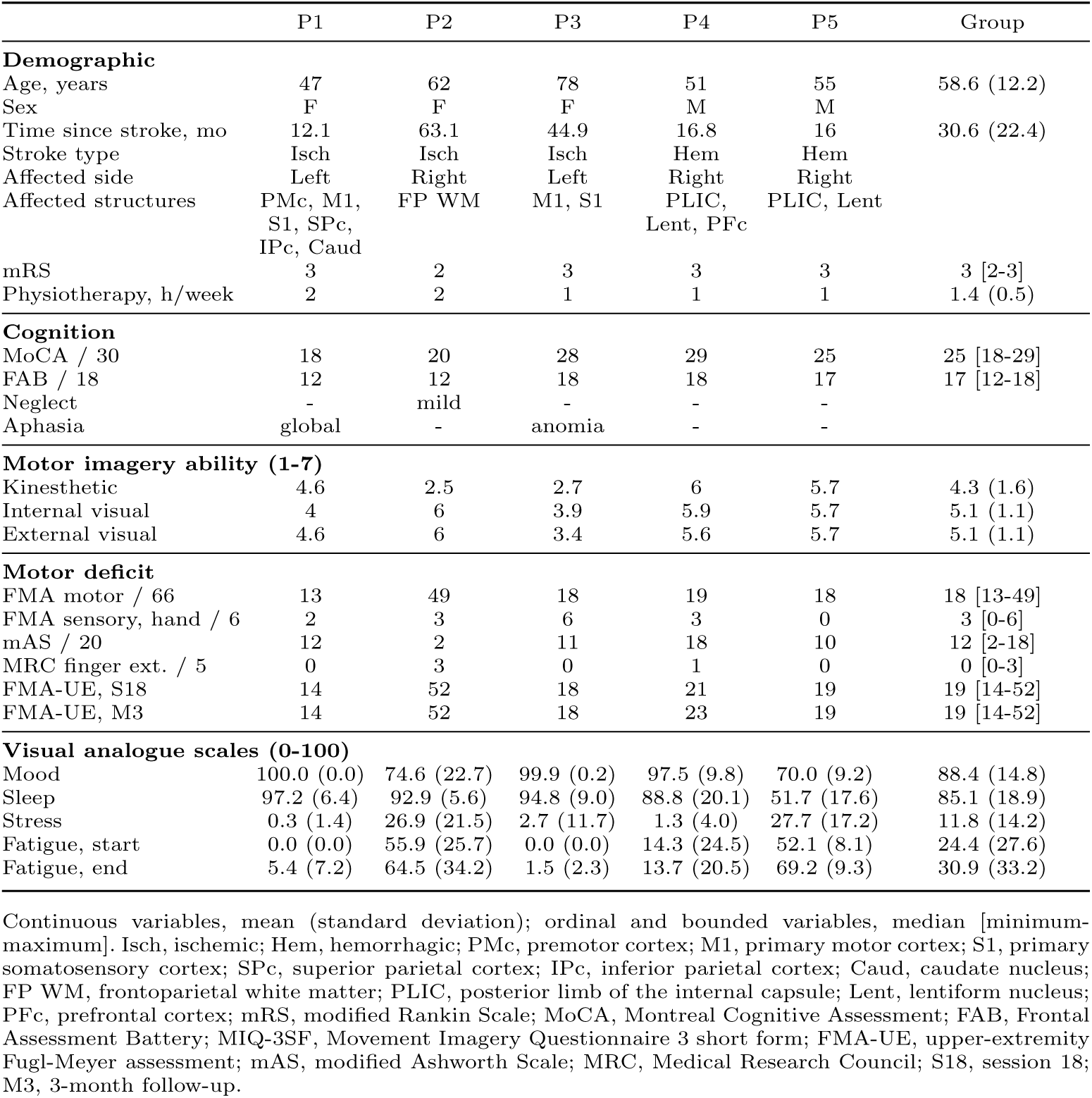
Participant characteristics and protocol measures.

At the group level, BCI control accuracy significantly improved across sessions, with the group mean increasing from 65.8% to 72.7% (ANCOVA, *p*_BH_ = 0.0015, **Fig. 3**). This result was confirmed offline by a progressive run-wise cross-validated accuracy increment indicating greater task separability (ANCOVA, *p <* 0.001, **Fig. S4a**). Consistently, rest distinctiveness increased across sessions, indicating that MI-related activity became progressively more distinct from rest (ANCOVA, *p <* 0.001, **Fig. S4b**). In contrast, class stability did not change over training, suggesting that improved BCI performance was not accompanied by a systematic reduction in the trial-to-trial dispersion of MI-related activity (ANCOVA, *p* = 0.42, **Fig. S4c**). Fatigue and spasticity changed significantly neither across sessions nor within sessions (ANCOVAs, *p >* 0.12, **Fig. S5**). Notably, we found no significant effects of the MI classification timing (MIT) on BCI accuracy, fatigue and spasticity (ANCOVAs, all *p ≥* 0.22).

**Fig. 3.**
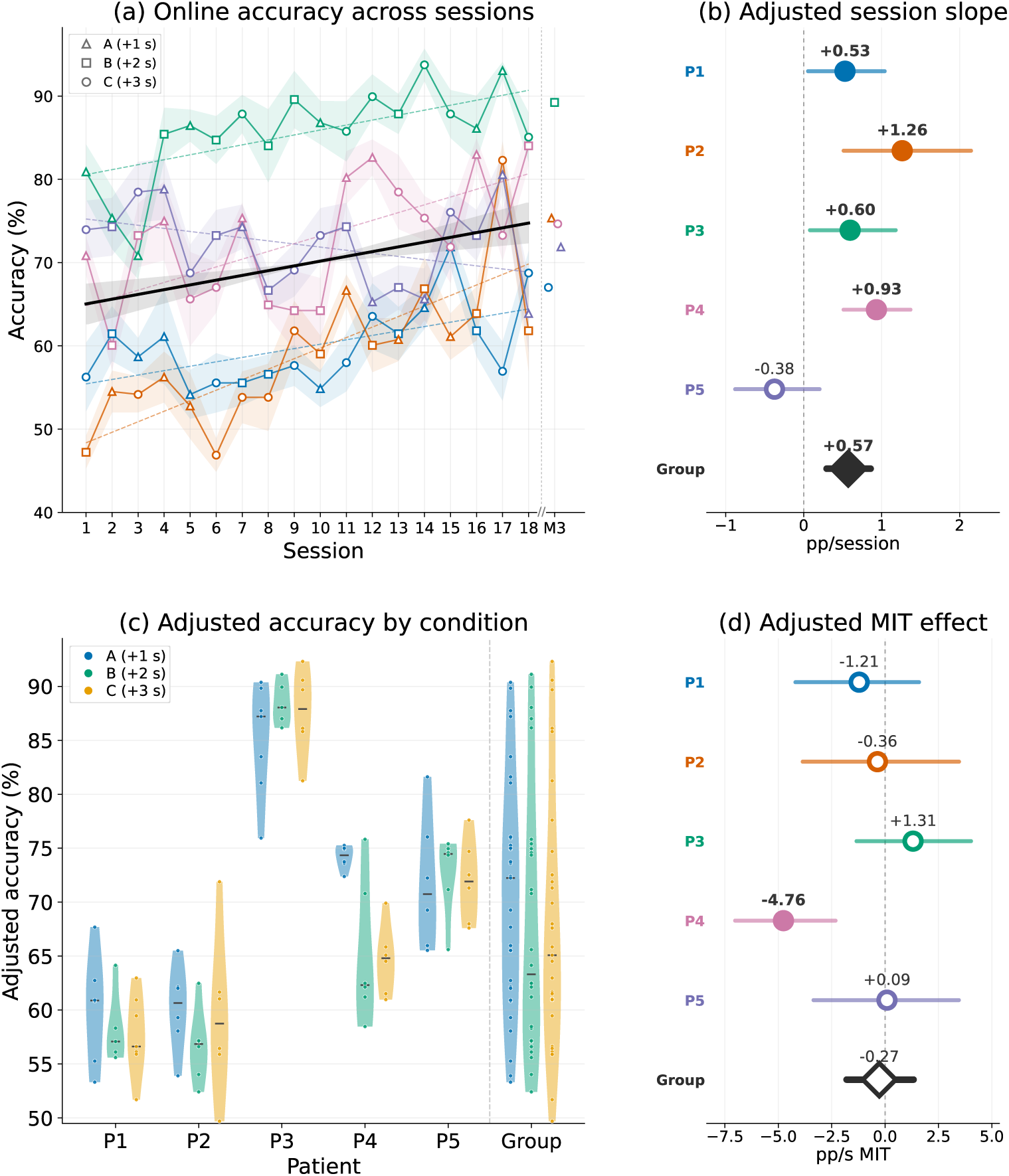
Online classification accuracy by session and MIT condition. (a) Individual trajectories across the 18 sessions. Shaded bands indicate the standard error across the eight runs, and marker shape denotes the MIT condition for each session (triangle, +1 s; square, +2 s; circle, +3 s after cue). For each patient, the colored dashed line shows the adjusted session trend, with the ANCOVA model slope reported in the forest plot in (b). The black line shows the group-level trend with its 95% confidence band. M3 is displayed beyond the dashed axis break for reference and is excluded from all statistical analyses. (c) Accuracy after adjustment for session and pretraining fatigue for the three MIT conditions; the pooled group distributions are shown separately to the right of the dashed vertical line (d) Forest plot of the adjusted MIT effect, expressed per additional second after cue. In both forest plots, the dashed vertical line indicates the null effect, circles denote patient-level estimates, and the diamond denotes the group estimate; filled markers indicate statistically significant effects.

At individual level, BCI accuracy improved significantly in four of the five patients (ANCOVAs, *p ≤* 0.034, **Fig. 3**). Offline, run-wise cross-validated accuracy increased in all five patients (ANCOVAs, all *p ≤* 0.022, **Fig. S4a**), while increases in rest distinctiveness were only clearly observed in P2, P3, and P4. No patient showed a clear longitudinal change in class stability (ANCOVAs, all *p≥* 0.14, **Fig. S4c**). No significant changes were found for fatigue or spasticity, except for an improvement of the within-session fatigue across sessions in P5 (ANCOVAs, *p* = 0.01, **Fig. S5**). Notably, two patients (P1, P3) reported no fatigue at all across the entire BCI intervention, and P2 had no spasticity. In general, the MIT did not have significant effects, but patient P4 exhibited deteriorated BCI accuracies when considering longer MIT (ANCOVA, *p* = 0.004).

### 3.2 MI-related spectral and spatial responses

Across the total 90 sessions, we found 126 significant clusters in the signed-*r*^2^ maps of the power spectra showing that MI values (*ϕ*) were in general lower than Rest (80 vs 46). Those values were predominantly observed in the beta band (95 vs 31 in alpha) and occurred more frequently over the unaffected hemisphere than the affected hemisphere (60 vs. 30) (**Fig. S6**). As for the node strength (*s*), we found 108 significant clusters in the respective signed-*r*^2^ maps, also indicating MI decrements compared to Rest (77 vs 31), with a less pronounced beta predominance (64 vs. 44) (**Fig. S7**). The mean absolute value *|r*^2^*|* ranged from 0.11 to 0.36 across patients for power spectrum and from 0.06 to 0.23 for node-strength, with the highest values observed in P3.

While, MI power decreases were predominant and spatially reproducible, the balance between negative (MI*<*Rest) and positive (MI*>*Rest) responses varied substantially across patients. P2, P4, and P5 predominantly exhibited MI-related negative responses, whereas P1 showed more positive responses. P3 consistently exhibited both negative and positive responses, with a focal beta power decrease at Cz accompanied by predominantly alpha power increase over bilateral fronto-central electrodes. The node strength values largely reproduced these patterns, although the positive responses were particularly sparse in P2, P4, and P5 (**Fig. S7**).

Spatial stability also differed markedly between patients. P3 showed the strongest spatial concentration, with beta negative responses localized at Cz in 15 of out 18 sessions. P5 showed a similarly concentrated distribution, although without convergence on a single electrode. P1 developed a stable positive response at C4 during the last sessions, whereas its negative response remained spatially distributed. P4 showed no clear spatial clustering and P2 had the most heterogeneous distribution particularly for positive responses (**Fig. S6**).

### 3.3 Evolution of ERD/ERS dynamics across sessions

The ERD/ERS temporal profiles corresponding to the strongest signed-*r*^2^ values further revealed marked inter-individual differences. ERDs emerged rapidly in P2 and P3, whereas they developed progressively in P4 and P5. In P1, a brief cue-related ERD was followed by a sustained ERS. P3 also showed a sustained ERS following cue onset. These profiles indicate substantial inter-individual variability in the temporal dynamics of MI-related brain activity (**Fig. 4**). Analyses of the amplitude, trial-wise standard deviation, duration and latency of ERD/ERS responses are presented in Table 2. In the MI classification windows, ERD amplitudes ranged from *−*14 to *−*56% (group mean *−*32%, SD 16), compared with +18 to +59% for ERS (group mean +32%, SD 19). ERD occupied 53 *−* 95% of the cue window (group mean 74%), compared with 13 *−* 75% for ERS (group mean 46%), and emerged earlier (590 vs 1100 ms after cue). Across sessions, ERD became stronger (*−*0.70%/session, *p*_BH_ = 0.018), less variable (*−*0.46, *p*_BH_ = 0.006), and more sustained within the MI phase (+1.6%/session, *p*_BH_ = 0.003), while its latency remained unchanged (*p*_BH_ = 0.29). ERS amplitude (+1.6%/session, *p*_BH_ = 0.018) and window occupancy (+1.9%/session, *p*_BH_ = 0.018) also increased, without significant changes in variability or latency. The Rest phase remained largely stable at the group level, with no significant session-related changes (all *p*_BH_ *≥* 0.09). Only P3 showed significant session-related changes during Rest, the window becoming progressively more occupied by an ERD (+3.4 pp/session, *p*_BH_ = 0.012), with no change in amplitude (**Fig. 4**).

**Fig. 4.**
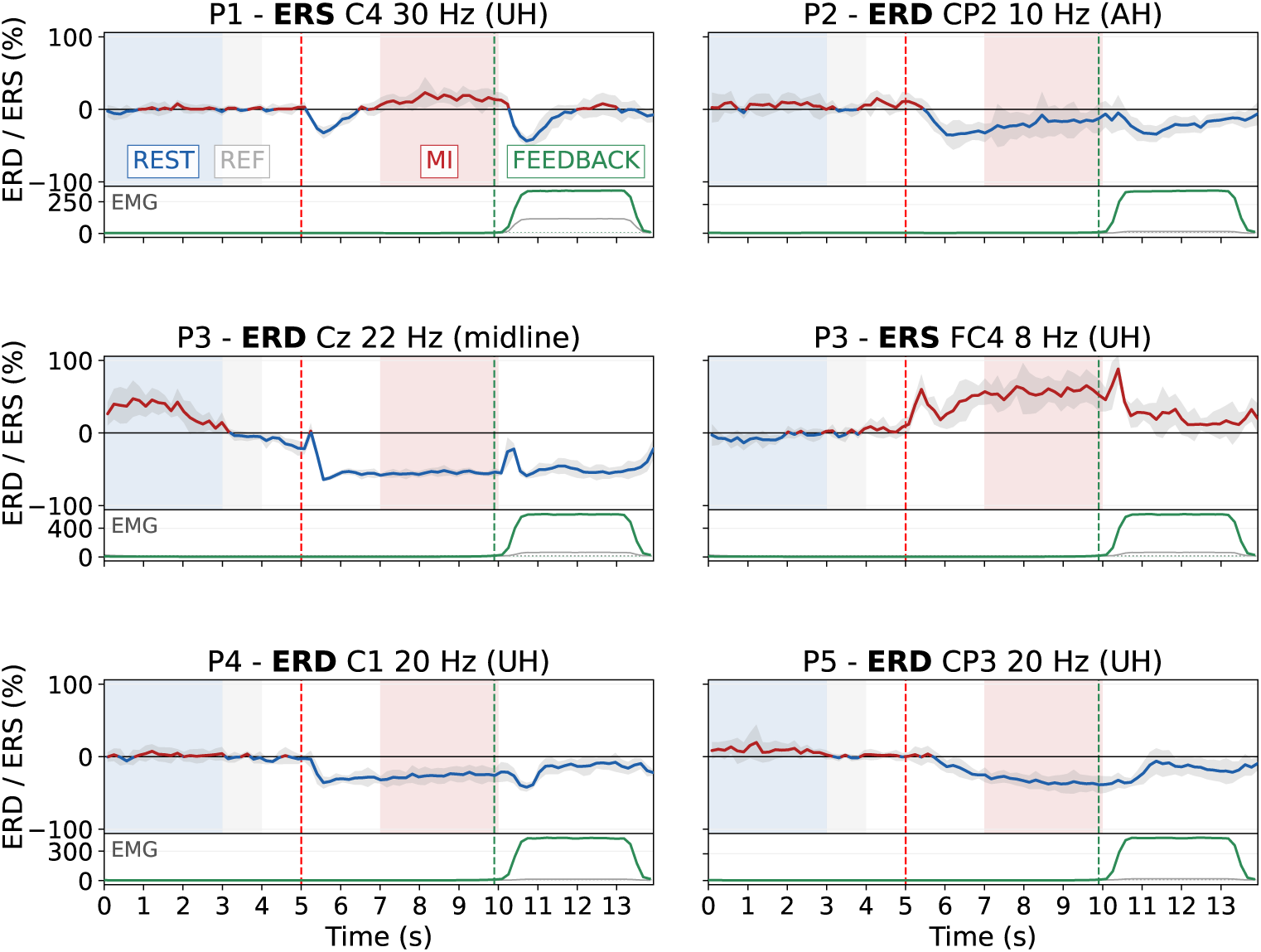
Session-averaged ERD/ERS time course and forearm EMG at a fixed electrode and frequency. Six panels for five patients: P3 contributes two sites, one ERD and one ERS. Only the +2 s MIT condition (B) is shown, so that imagery duration is matched across patients. Relative PSD change below and above the reference (3-4 s, grey band) is shown as ERD (blue trace) and ERS (red trace). Shading is the standard deviation across sessions. Blue and red shaded bands mark the online rest and MI analysis windows. Lower sub-panels show the forearm EMG envelope, paretic side in green and non-stimulated side in grey, on the same time axis. The dashed red line marks the cue; the dashed green line marks the FES trigger, in average *−*102 ms before the end of the online MI analysis window. AH, affected hemisphere; UH, unaffected.

**Table 2.** Session-wise evolution of the trial outcomes and effect of motor imagery time. Effects are given in each patient and at the group level.

|  | **P1** |  | **P2** |  | **P3** |  | **P4** | **P5** |  | **Group** |  |
|---|---|---|---|---|---|---|---|---|---|---|---|
|  | neg. | pos. | neg. | pos. | neg. | pos. | neg. | neg. | pos. | neg. | pos. |
| *n* | 14 | 18 | 13 | 8 | 18 | 18 | 17 | 18 | 2 | 80 | 46 |
| **3-s Rest window** |  |  |  |  |  |  |  |  |  |  |  |
| *AUC*, % change |  |  |  |  |  |  |  |  |  |  |  |
| Mean | 7.6 | -4.6 | 5.3 | -5.5 | 27 | -3.5 | 6 | 9.2 | 1.5 | 11 | -3.0 |
| (SD) | (12) | (5.1) | (9.5) | (5.7) | (14) | (5.8) | (7.7) | (10) | (13) | (9) | (3.1) |
| Session | -1.4 | 0.27 | -1.3 | -0.35 | 1.4<sup>†</sup> | 0.53<sup>†</sup> | 0.027 | 0.34 | - | 0.14 | 0.39<sup>†</sup> |
| MIT | -2.5 | -0.35 | -1.4 | -3.3 | 3.6 | 1.3 | 3.1 | 0.055 | - | 1.1 | -0.26 |
| *Duration*, % of windows |  |  |  |  |  |  |  |  |  |  |  |
| Mean | 12 | 11 | 14 | 19 | 73 | 6.8 | 16 | 23 | 5.6 | 28 | 11 |
| (SD) | (23) | (16) | (19) | (21) | (27) | (7.3) | (16) | (27) | (7.9) | (26) | (6.3) |
| Session | -1.3 | -0.94 | -0.35 | 1.4 | **3.4** | **-0.9** | 0.25 | 0.56 | - | 0.85 | -0.69 |
| MIT | -9.5 | 0.56 | 1.7 | 11 | 7 | -2.4 | 1.4 | 2.1 | - | 2.2 | 0.75 |
| **3-s MI window** |  |  |  |  |  |  |  |  |  |  |  |
| *AUC*, % change |  |  |  |  |  |  |  |  |  |  |  |
| Mean | -14 | 30 | -21 | 20 | -56 | 59 | -35 | -31 | 18 | -32 | 32 |
| (SD) | (9.3) | (16) | (17) | (16) | (7.1) | (30) | (9.1) | (8.1) | (5.9) | (16) | (19) |
| Session | -0.98 | 0.87 | **-2.5** | 0.19 | -0.099 | **2.9** | -1.2<sup>†</sup> | 0.42 | - | **-0.70** | **1.6** |
| MIT | 1 | -5.5 | -2 | 9.3 | 0.43 | 12 | 4.2 | -0.71 | - | 0.46 | 5.3 |
| *Between-trial SD*, % change |  |  |  |  |  |  |  |  |  |  |  |
| Mean | 31 | 46 | 30 | 44 | 14 | 62 | 20 | 20 | 49 | 23 | 50 |
| (SD) | (10) | (12) | (8.5) | (23) | (1.7) | (18) | (3.4) | (5.4) | (7.2) | (7.4) | (7.9) |
| Session | **-1.4** | 0.40 | **-1.4** | -1.3 | 0.034 | 1.3 | -0.43<sup>†</sup> | 0.15 | - | **-0.46** | 0.35 |
| MIT | 4.9 | -5.9 | 0.077 | 16 | -1 | -0.27 | 1.3 | 1.5 | - | 1 | 1.1 |
| **Cue time** |  |  |  |  |  |  |  |  |  |  |  |
| *Duration*, % of windows |  |  |  |  |  |  |  |  |  |  |  |
| Mean | 53 | 61 | 53 | 35 | 95 | 75 | 90 | 78 | 13 | 74 | 46 |
| (SD) | (27) | (22) | (32) | (25) | (3.2) | (19) | (6) | (9.3) | (0.43) | (20) | (28) |
| Session | **4.7** | **2.9** | **5.4** | 1.9 | 0.36<sup>†</sup> | 0.59 | 0.48 | -0.37 | - | **1.6** | **1.9** |
| MIT | -10 | -8.9 | -1.7 | 8.3 | -0.49 | 8.7 | -1.2 | -1.9 | - | -2.6 | 0.53 |
| *Latency*, ms |  |  |  |  |  |  |  |  |  |  |  |
| Mean | 446 | 420 | 893 | 2093 | 376 | 463 | 331 | 900 | 1449 | 589 | 1106 |
| (SD) | (373) | (530) | (325) | (1262) | (80) | (394) | (89) | (360) | (1594) | (283) | (811) |
| Session | **-63** | -44 | -27 | -72 | -9.3 | 18 | -0.38 | 13 | - | -12 | -43 |
| MIT | -70 | 0.68 | 199 | 744 | 53 | -276<sup>†</sup> | 23 | 120 | - | 39 | 116 |

neg., negative cluster peak; pos., positive cluster peak. *n*, sessions carrying a cluster, out of 18 per patient and pooled in the Group columns. Session, slope per session; MIT, motor imagery time, per additional second beyond 4 s. Mean and SD are taken across sessions, and across the patient means in the Group columns. P4 carried no positive cluster peak, and P5 carried one in two sessions only, too few for a slope estimation. Within every column, the group columns included, each effect is corrected by Benjamini-Hochberg over the six outcomes of that column: bold survives it, <sup>†</sup> is significant uncorrected only. -, not estimable.

Finally, MIT had significant effects on the longitudinal reinforcement of ERD and ERS neither at the group nor at individual level (group *p*_BH_ *≥* 0.59; individual *p*_BH_ *≥* 0.18).

### 3.4 Topography of the reinforcement and network coupling

Longitudinal reinforcement was spatially localized. Across sessions, every patient exhibited at least one significantly reinforced *|r*^2^*|* cluster in the power spectra (*ϕ*), negative in four of them and positive in P1, while all but one showed at least one such cluster in node strength (*s*) (**Fig. 5**).

**Fig. 5.**
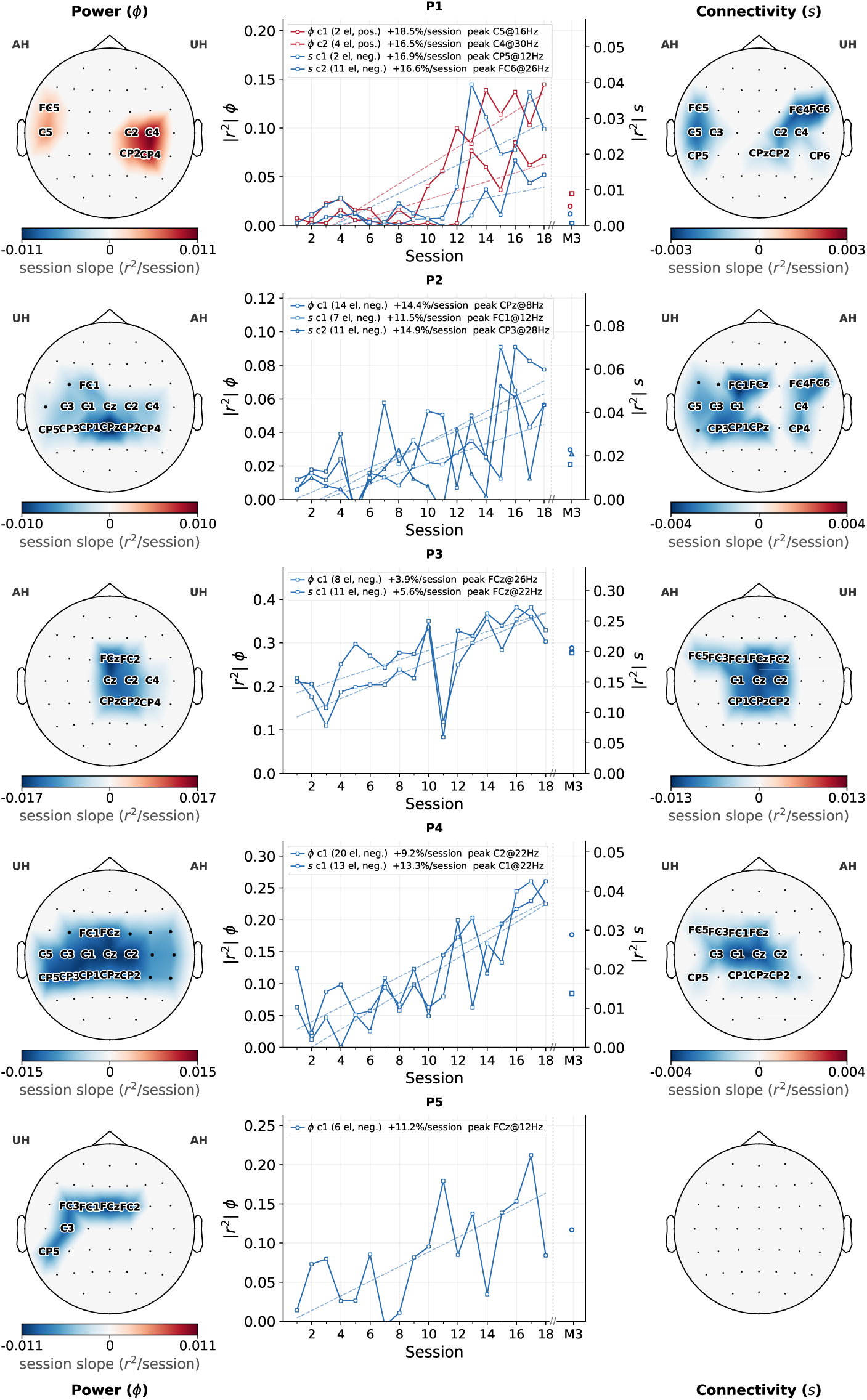
Longitudinal reinforcement of the power spectral density (*ϕ*) and node strength connectivity (*s*), one row per patient. Topographies show significant cluster slopes (black dots; twelve steepest electrodes labeled), with *ϕ* on the left and *s* on the right. Blue and red indicate decreasing and increasing signed *r*^2^, respectively; intensity reflects the rate of change. The central panel shows cluster-mean *|r*^2^| adjusted for imagery duration and baseline fatigue, with dashed fitted trends (left axis, *ϕ*; right axis, *s*). The unadjusted 3-month follow-up appears after the axis break and is excluded from the fit. Legends indicate cluster size, response sign, relative session slope, and peak electrode and frequency. AH, affected hemisphere; UH, unaffected hemisphere; *ϕ*, power spectral density; *s*, node strength.

The cluster slopes ranged from +3.9 to +18.5% across sessions. Since the slopes were calculated relative to each cluster’s polarity, positive slopes indicate that MI-Rest discriminability increased over time for both negative and positive responses (**Fig. 5**). MIT had no significant effect in four of the five patients, whereas in P4 a longer MIT was associated with reduced MI-Rest discriminability for both spectral power over frontocentral electrodes (12-26 Hz, *p* = 0.013) and node strength over central midline electrodes (8-10 Hz, *p* = 0.026), consistent with the decline in online accuracy observed in this patient (**Supplementary Tables, (Table S1)**).

Network analysis further revealed a consistent group-level decrease of the functional connectivity contrast in sensorimotor networks (**Fig. 6**). Inner motor edges showed a progressive decrease of the within-hemisphere segregation in both the affected and the unaffected hemisphere (*p*_BH_ *≤* 0.005), and a similar decrement for the between-hemisphere integration (*p*_BH_ = 0.002). The segregation decrement was larger in the affected hemisphere, as indicated by the laterality index, although this difference did not survive correction (*p*_BH_ = 0.057). Decomposing the contrast showed that these effects were driven by longitudinal increases in Rest connectivity rather than by decreases during MI, the MI component remaining flat in all three scopes (uncorrected, Rest *p ≤* 0.065 against MI *p ≥* 0.47; **Fig. S8**).

**Fig. 6.**
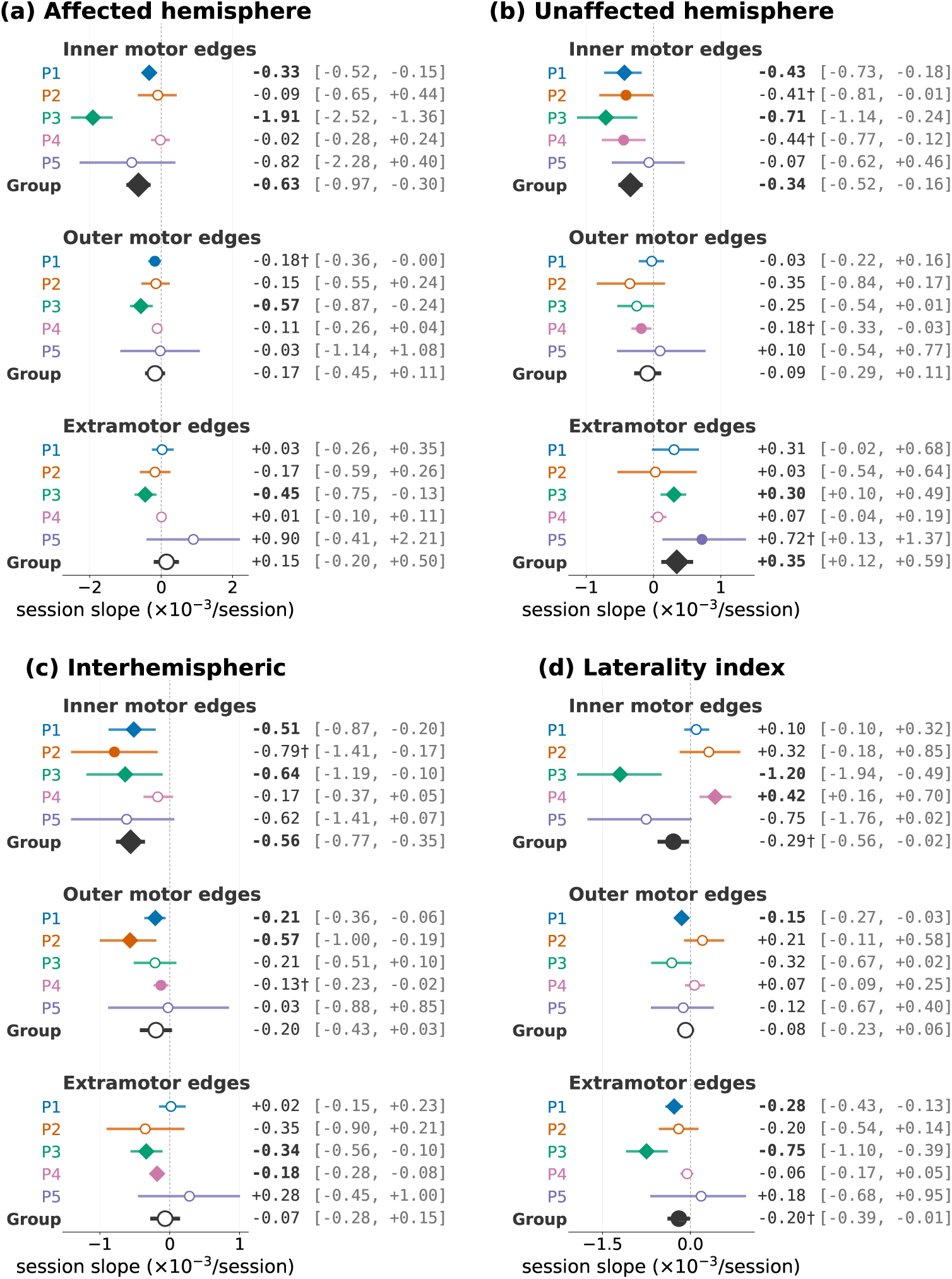
Session slope of network intensities, in 10^−3^ per session. Network intensity slopes across sessions, with one panel per network scope: (a) affected-hemisphere segregation, (b) unaffected-hemisphere segregation, (c) interhemispheric integration, and (d) laterality index, defined as the difference between (a) and (b). Each panel distinguishes inner motor, outer motor, and extramotor edges. Colors identify patients P1–P5, and black denotes the group estimate. Horizontal bars indicate 95% confidence intervals; filled circles indicate uncorrected significance (*p <* 0.05), and diamonds indicate significance after Benjamini–Hochberg correction. Corresponding estimates are marked with a dagger or boldface, respectively. The dashed vertical line indicates a null slope; negative values indicate decreasing network intensity across sessions.

No significant group effects were found for outer-motor edges (*p*_BH_ *≥* 0.17), and among extra-motor edges only the segregation of the unaffected hemisphere increased (*p*_BH_ = 0.006); the laterality index did not survive correction (*p*_BH_ = 0.057).

At the individual level, the connectivity trajectories were quite heterogeneous. P1 and P3 showed the most extensive decrements for inner motor edges, across all three scopes (P1, all *p*_BH_ = 0.009; P3, all *p*_BH_ *≤* 0.03), although the relative task contributions might differ: P1’s effects were primarily driven by increases in the Rest phase, whereas P3’s were predominantly driven by decreases during MI (**Fig. S8**). P2 showed a limited interhemispheric decrease for the outer motor edges (*p*_BH_ = 0.040), while P4 showed a shift in the inner motor hemispheric balance (*p*_BH_ = 0.015) and interhemispheric extramotor decoupling (*p*_BH_ = 0.013). No significant longitudinal connectivity effects were reported for P5 (all *p*_BH_ *≥* 0.18).

## 4 Discussion

### 4.1 Individual variability and BCI learning

This study identified three main features of repeated MI-BCI training in chronic stroke. First, BCI control changed across sessions, with marked differences in the magnitude and neurophysiological expression of these trajectories between patients. Second, varying the onset of the MI decoding window produced no systematic benefit within the tested range. Third, longitudinal changes in BCI control were accompanied by changes in both local spectral activity and functional connectivity, but without meaningful short-term motor improvement. Together, these results support an individualized and longitudinal interpretation of BCI control rather than a model based on a fixed neural response or a single temporal parameter. Such heterogeneity is consistent with previous reports of individual differences in MI-related oscillatory responses and their evolution after stroke [3, 4], including trajectories that diverge within the same cohort, recovering and non-recovering patients showing opposite patterns [5].

All patients but one showed increasing BCI accuracy across sessions, and a similar longitudinal trajectory was reproduced offline using run-wise cross-validated accuracy. Classifier-independent covariance measures clarified the nature of this longitudinal change. MI and Rest became progressively more separable, whereas the global within-class stability of MI did not increase. Improved BCI control therefore reflected increasing differentiation between MI and Rest rather than convergence toward a more stereotyped MI state. This means that improved BCI control was associated primarily with increasing MI-rest separability rather than with convergence toward a more stereotyped global MI state. The convergence of online control, offline decodability, and neurophysiological changes supports an interpretation in terms of acquisition of BCI control, although the observational progression across sessions does not by itself establish learning as the causal mechanism. This distinction is relevant for restorative BCI rehabilitation because successful classification alone does not establish that a recovery-relevant neural state is being reinforced. The present results identify physiological changes associated with acquisition of BCI control, but do not establish whether these changes are themselves restorative. This is important because MI-BCI training is commonly framed around reinforcement of a canonical ERD response [13, 46, 47], yet we showed that the MI-related brain activity and connectivity responses strengthened by the BCI training were not uniform across patients.

In general, ERD is typically associated with sensorimotor activation [36], whereas the post-movement beta rebound has been interpreted as an idling state of primary motor cortex [48], and surrounding ERS as inhibition or disengagement of nonrecruited motor areas [49]. Beta rebounds may also reflect confidence in internal-model estimates rather than idling alone [50]. In P1, the effective feedback signal was associated with a beta rebound, whereas P3 received feedback associated with an ERD. Both showed longitudinal strengthening of ERS-related activity, possibly due to the proprioceptive feedback associated with passive movements and tactile stimulation [51, 52] (**Fig. 4**). These findings suggest that BCI learning in stroke rehabilitation should not be understood as the progressive stabilization of a single canonical MI pattern. Personalization should consequently extend beyond classifier calibration to the selection and longitudinal reassessment of the brain features used for feedback.

### 4.2 MI timing and temporal dynamics

Allowing the classifier to start the MI decoding longer after the cue did not systematically improve BCI accuracy or longitudinal neural outcomes. Although late classification may accommodate slower or altered MI responses after stroke [15, 53], our results indicate that this is unlikely to benefit all patients.

The ERD/ERS temporal profiles observed here illustrate why this distinction matters (**Fig. 4**). In P1, all the three possible MI classification windows captured a sustained beta rebound because the preceding ERD had already ended. More in general, a fixed decoding window may mismatch the temporal dynamics of an individual’s MI response even when the underlying response is robust. This has implications for the interpretation of the MI timing effects. The absence of a group-level effect of MI timing does not necessarily imply that timing is irrelevant. Rather, its effect may be obscured by heterogeneous individual response dynamics. A MI classification timing that is appropriate for one participant may be suboptimal for another, particularly when the onsets and durations of ERD/ERS differ.

The timing of the decoding window should therefore be considered in relation to the temporal dynamics of the individual MI response rather than treated as a universally optimal parameter. Future protocols could align the decoding window with each patient’s MI response profile and reassess this alignment as the response evolves across training.

### 4.3 Clinical and network changes during training

Despite longitudinal changes in BCI control, brain activity, and functional connectivity, the intervention was not accompanied by meaningful short-term motor improvement. Patients gained between 0 and 3 points on the UE-FMA, remaining below the reported clinically important difference of 4.25 to 7.25 points [54]. The present results therefore provide no evidence that the BCI protocol used in this pilot improved upper-limb motor function.

The BCI intervention was not systematically embedded within a standardized program of concurrent physiotherapy or occupational therapy. Such combined rehabilitation may be important for translating BCI-related neural changes into repeated task-specific motor practice. In addition, several participants had very severe distal impairment, and eighteen sessions may represent a relatively short exposure for detecting motor change in this population [6].

Functional connectivity provided complementary information to local spectral activity. Node strength also showed longitudinal changes over sensorimotor regions in four of five patients, indicating progressive changes in functional coupling between MI and Rest [20, 21, 55]. These effects should not be interpreted as a uniform increase or decrease in beneficial network engagement, because node strength quantified a task contrast rather than connectivity during MI alone.

At the group level, edge-level analysis showed a progressive decrease in within-hemisphere and interhemispheric coupling for connections within the motor network. These patterns are compatible with the view of stroke as a network-level disorder in which reorganization can involve ipsilesional re-engagement, contralesional compensation, and interhemispheric redistribution [16–18, 32, 56–58].

However, these connectivity changes quantified the MI-Rest contrasts, so an over-all decrease can result from connectivity decrease during MI, increase during Rest, or both. For example, in patients P1 and P4 connectivity decrements were predominantly modulated by Rest increases, consistent with evolving baseline network reorganization [4, 59]. Instead, P3 showed primarily pure MI-driven modulations, consistent with task-related reorganization [46, 47]. Analyzing MI and Rest separately is therefore essential when interpreting longitudinal connectivity changes and dedicated resting-state recordings would be required to determine whether changes observed during the reference period reflect broader changes in ongoing network organization.

Together, these findings suggest that BCI learning in stroke patients extends beyond the local spectral features. Spectral power captures local oscillatory modulation, whereas functional connectivity provides information about interactions across regions. The associated trajectories may therefore provide complementary markers of BCI learning, although whether they predict longer-term functional recovery remains to be elucidated.

### 4.4 Implications for personalized BCI rehabilitation

These converging findings support treating personalization as a longitudinal process. During BCI training, the patient learns to generate a detectable signal, the decoder learns to recognize it, and the underlying brain activity and network organization may themselves change [60]. These two learning processes are coupled, and the underlying brain activity and network organization may themselves evolve across training. Online BCI accuracy alone cannot distinguish these processes [35].

Three aspects emerge from these results as relevant to longitudinal personalization. First, the physiological feature used for feedback may need to be selected according to the patient’s actual MI response rather than assuming a canonical ERD. Second, the timing of the decoding window may need to reflect the onset and duration of the individual’s response and potentially adapt as training progresses. Third, brain connectivity measures may complement local spectral features when characterizing changes that are not captured by the classifier alone.

This perspective also reframes the interpretation of BCI learning. Improved classification does not necessarily indicate convergence toward a common neural mechanism, just as a stable clinical score does not imply an absence of neural adaptation. Instead, behavioral, spectral, and network trajectories may provide partially independent views of the learning process. Identifying which of these trajectories are associated with later functional recovery is therefore an important open question.

The present study does not establish which brain feature or trajectory is optimal or causally related to recovery. Its main contribution is instead to show that repeated BCI training can produce individual changes in the neural signals used for control that are not fully captured by a single group-level feature or fixed protocol parameter. Larger controlled studies should determine whether these trajectories identify distinct BCI-learning phenotypes, how BCI parameters can be adapted during training, and eventually whether neural trajectories predict longer-term clinical recovery.

### 4.5 Conclusion

Longitudinal MI-BCI training in chronic stroke was accompanied by measurable changes in control accuracy, brain activity, and functional connectivity despite the absence of meaningful short-term clinical improvement. Negative responses (MI*<*Rest) associated with ERDs were generally predominant, although positive responses (MI*>*Rest) could also become robust and behaviorally useful signals. These findings challenge the assumption that a single MI response or fixed temporal parameter is appropriate for all patients. Personalization should instead be viewed as a longitudinal process in which the relevant brain activity and connectivity, as well as their timing, may evolve with training. Whether these trajectories can be used to adapt BCI parameters and predict longer-term clinical recovery remains perhaps the most important open question to address in future research.

## Data Availability

All data produced in the present work are contained in the manuscript

## Declarations

### Consent for publication

Not applicable.

### Availability of data and materials

The datasets analyzed during the current study are not publicly available due to patient confidentiality. They are available from the corresponding author on reasonable request, subject to approval by the study sponsor and a data sharing agreement in accordance with French regulations.

### Authors’ contributions

CB: Conceptualization, Methodology, Investigation, Data curation, Validation, Formal analysis, Visualization, Writing - original draft, Supervision, Project administration, Software. CG: Investigation, Data curation, Validation, Formal analysis. DB: Investigation. ES: Investigation, Data curation. MC: Investigation, Data curation. GA: Investigation, Resources. MO: Investigation, Resources. CZ: Investigation, Resources. SD: Investigation, Resources. EB: Investigation, Resources. LB: Methodology, Supervision. PB: Supervision, Project administration, Resources. FDVF: Methodology, Supervision, Project administration, Funding acquisition, Resources. All authors contributed to writing - review & editing, and read and approved the final manuscript.

## Acknowledgements

Not applicable.

