## Supplementary material for "Single-Case Design for Network-based BCI Rehabilitation: Individual Learning Trajectories and Effects of Motor Imagery Timing in Stroke Patients": Supp. Figures

### Supplementary Figures

Supplementary material to: *A single-case approach to BCI protocol design: individual learning trajectories and the calibration of motor imagery time after stroke.*

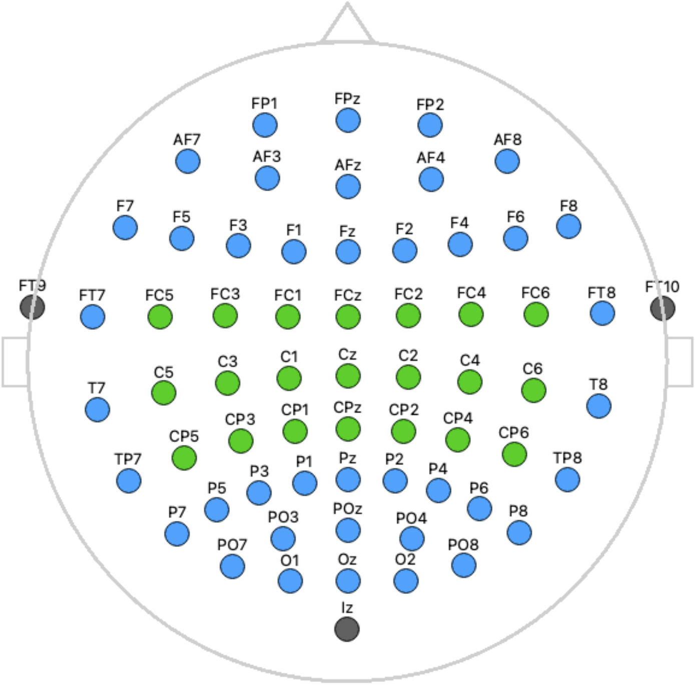


**Fig. S1 Electrode montage.** Scalp topography of the 64-electrode cap, arranged according to the extended 10-20 system. Electrodes highlighted in green (FC, C, and CP rows) denote the sensorimotor region of interest used throughout the analyses. Electrodes shown in grey (FT9, FT10, and Iz) were excluded from all analyses.

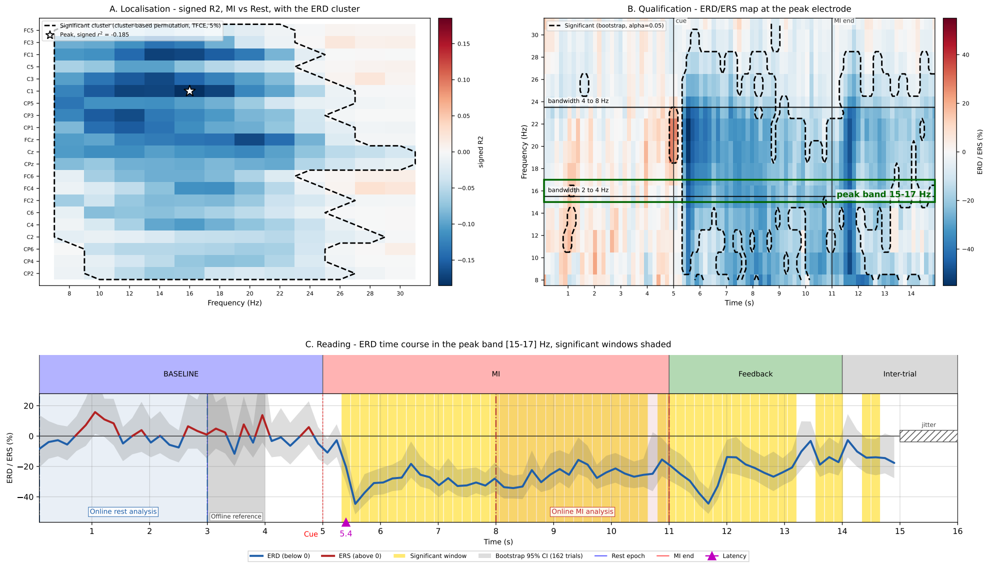


**Fig. S2 Extraction and temporal characterization of MI-related ERD/ERS responses.** Illustration of the analysis procedure for one representative ERD response. **(A)** The MI-Rest signed $r^2$ map was used to identify significant channel-frequency clusters within the sensorimotor electrodes; the star indicates the peak of the selected cluster. **(B)** At the peak electrode, the time-frequency map of relative power change was used to characterize the temporal extent of the response. Dashed contours indicate windows significantly different from baseline, and the green lines delimit the frequency band containing the cluster peak. **(C)** Time course of the relative power change averaged within the peak frequency band. ERD and ERS are shown below and above zero, respectively; yellow shading denotes significant windows and grey shading the bootstrap 95% confidence interval across trials. The online Rest and MI analysis windows, offline reference period, cue onset, end of the MI period, and estimated response latency are indicated.

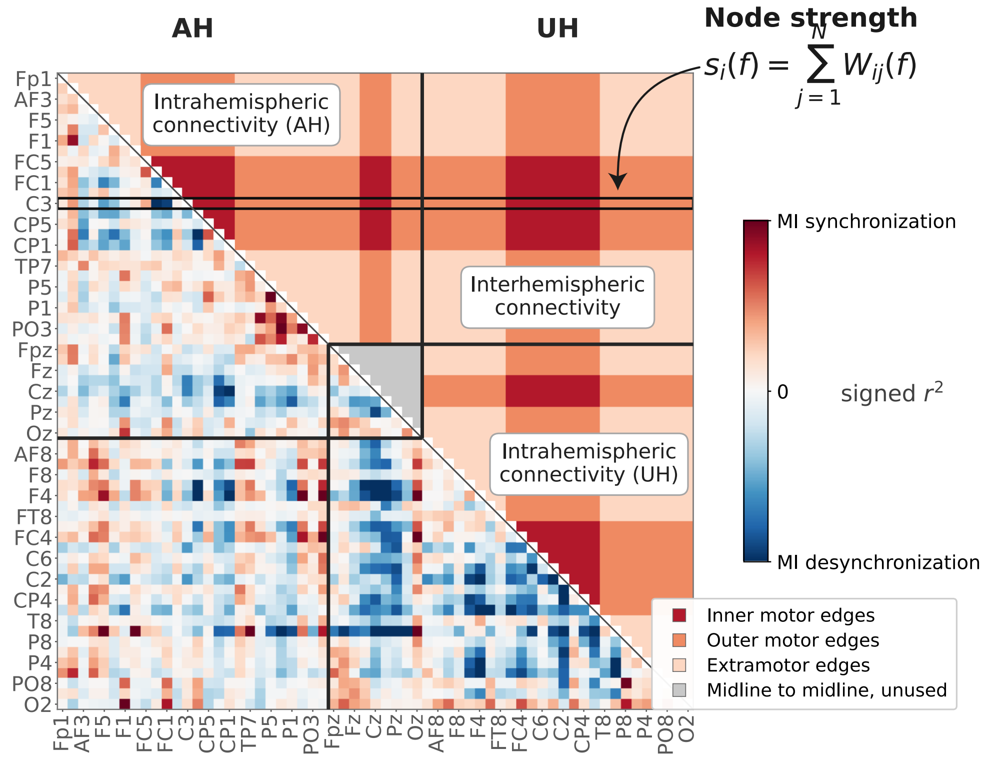


**Fig. S3 Network measures and edge partitioning.** Schematic representation of node-strength and edge-level MI-Rest connectivity analyses. Edges were classified according to their hemispheric contribution (affected hemisphere, AH; unaffected hemisphere, UH; or interhemispheric) and their relation to the motor network. Inner motor edges connected two motor electrodes ($n = 210$: 63 within AH, 63 within UH, 81 interhemispheric), outer motor edges connected one motor and one non-motor electrode ($n = 840$: 258, 258, and 306, respectively), and extramotor edges connected two non-motor electrodes ($n = 780$: 238, 238, and 289, respectively). The remaining 36 midline-to-midline edges were not assigned to a hemisphere and were excluded from hemispheric analyses. Signed $r^2$ values indicate relative MI increased connectivity (positive) or decreased connectivity (negative) compared with Rest.

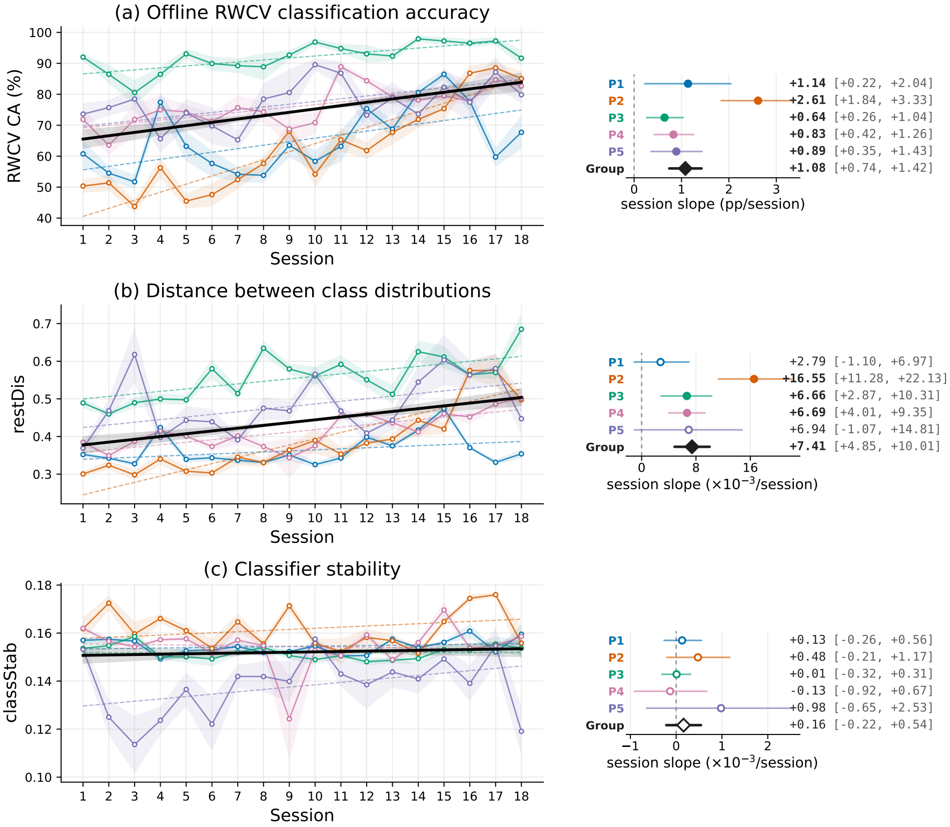


**Fig. S4 Offline measures of BCI learning across sessions.** **(a)** Run-wise cross-validated classification (RWCV) accuracy, quantifying offline MI-Rest decodability with a constant amount of training data. **(b)** Rest distinctiveness, quantifying the separation between the MI and Rest covariance distributions relative to their dispersion. **(c)** MI-class stability, quantifying the within-class consistency of MI covariance patterns. Colored trajectories represent individual patients and the black line, the group-level session trend; shaded bands indicate variability across runs. Forest plots show the adjusted session slopes with 95% confidence intervals, with diamonds indicating group estimates. Filled markers denote statistically significant session effects.

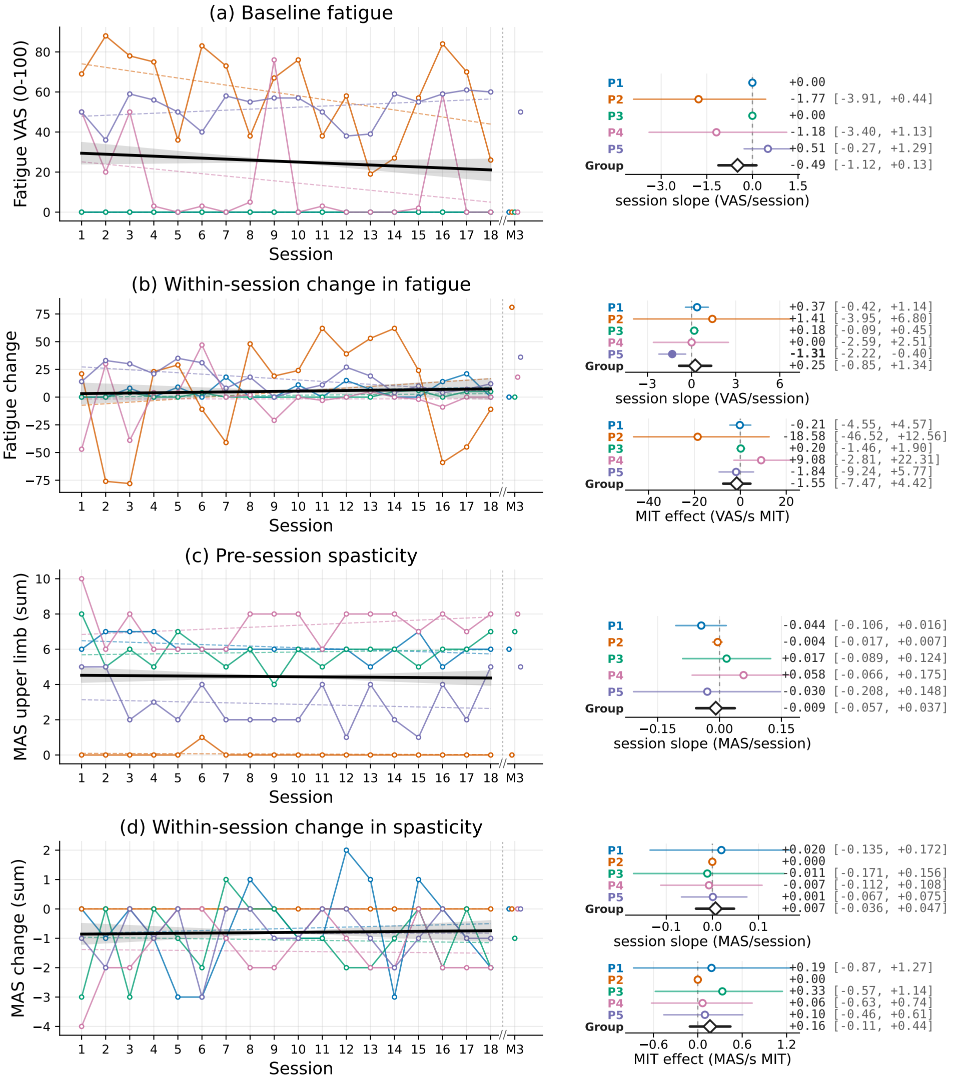


**Fig. S5 Evolution of fatigue and spasticity across BCI training. (a)** Pre-session fatigue and **(b)** within-session change in fatigue across the 18 training sessions. **(c)** Pre-session upper-limb spasticity and **(d)** within-session change in spasticity. Colored trajectories represent individual patients and the black line, the group-level session trend. Forest plots show the adjusted session slopes and, for within-session changes, the effect of motor imagery classification timing (MIT) per additional second. Horizontal bars indicate 95% confidence intervals and diamonds, the group estimates. The 3-month follow-up (M3) is displayed after the axis break for reference and was excluded from the longitudinal analyses. VAS, visual analogue scale; mAS, modified Ashworth Scale.

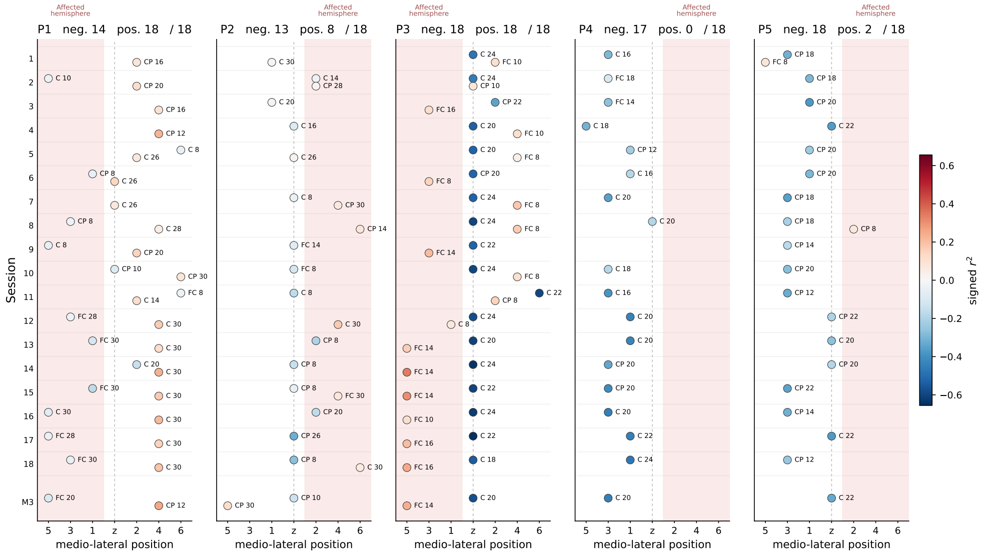


**Fig. S6 Spatial distribution of session-wise spectral $r^2$ peaks.** For each patient and session, the most discriminant significant spectral-power peak was identified separately for negative $r^2$, blue and positive $r^2$, red within sensorimotor electrodes and the 8-30 Hz range. Each marker is positioned according to the medio-lateral location of the corresponding electrode and labeled with the electrode name and peak frequency (Hz). The shaded region indicates the affected hemisphere, and the dashed vertical line the midline. Numbers above each panel indicate the number of sessions containing a significant positive or negative peak out of 18. M3 denotes the 3-month follow-up and was not included in the longitudinal analyses.

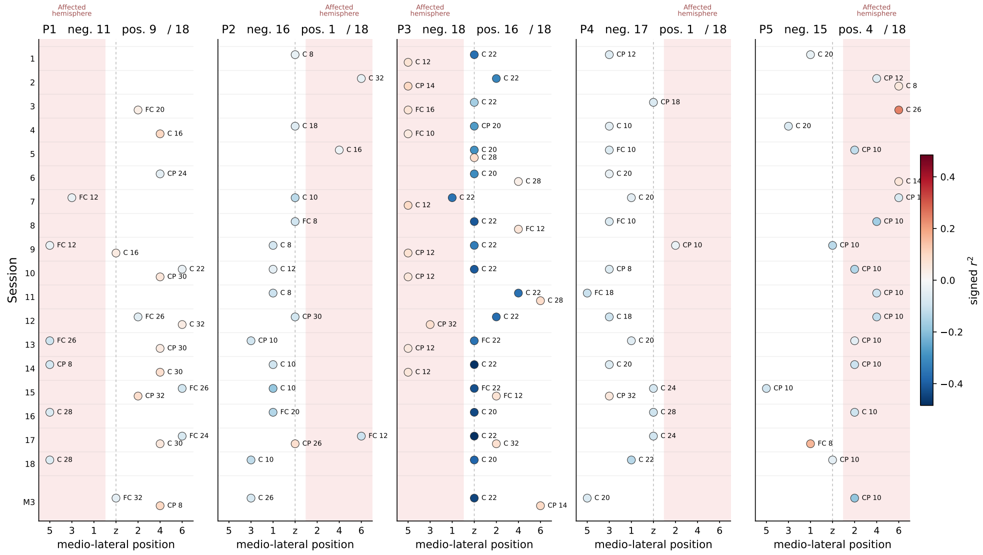


**Fig. S7 Spatial distribution of session-wise node-strength negative and positive peaks.** For each patient and session, the most discriminant significant node-strength peak was identified separately for negative $r^2$, blue and positive $r^2$, red within sensorimotor electrodes and the 8-30 Hz range. Each marker is positioned according to the medio-lateral location of the corresponding electrode and labeled with the electrode name and peak frequency (Hz). The shaded region indicates the affected hemisphere, and the dashed vertical line the midline. Numbers above each panel indicate the number of sessions containing a significant negative or positive peak out of 18. M3 denotes the 3-month follow-up and was not included in the longitudinal analyses.

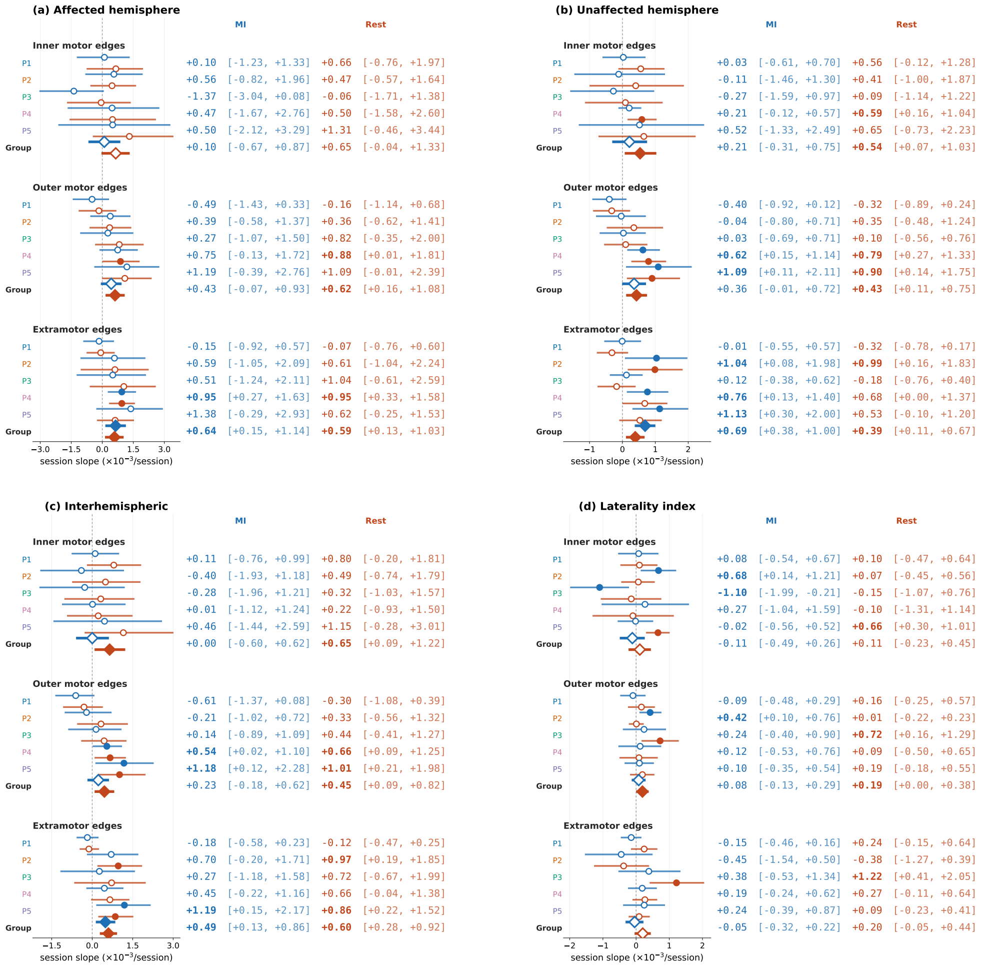


**Fig. S8 MI and Rest contributions to longitudinal connectivity changes.** Session slopes of connectivity analyzed separately during MI and Rest for **(a)** the affected hemisphere, **(b)** the unaffected hemisphere, **(c)** interhemispheric connections, and **(d)** the laterality index. Within each compartment, edges are separated into inner motor, outer motor, and extramotor connections. Individual patient estimates are shown in color, while group estimates are indicated by diamonds and reported separately for MI and Rest. Horizontal bars indicate 95% confidence intervals and the dashed vertical line the null slope. Comparing MI and Rest slopes identifies whether longitudinal changes in the MI-Rest connectivity contrast were predominantly driven by changes during imagery, rest, or both.
