## Supplementary material for "Single-Case Design for Network-based BCI Rehabilitation: Individual Learning Trajectories and Effects of Motor Imagery Timing in Stroke Patients": Supp Methods

### Supplementary Methods

Supplementary material to: *A single-case approach to BCI protocol design: individual learning trajectories and the calibration of motor imagery time after stroke.*

#### S1 Eligibility and inclusion assessment

##### S1.1 Inclusion criteria

Patients were eligible if they had sustained a single stroke more than six months before the inclusion visit; presented a distal upper-limb motor deficit, defined as an upper-extremity Fugl-Meyer score below 53 [1] with visible active finger flexion (Medical Research Council grade 2 or above) [2]; were right-handed; were aged between 18 and 85 years; had given written informed consent; and were affiliated to a social security scheme.

##### S1.2 Non-inclusion criteria

Patients were not included if they were under legal protection or deprived of liberty, pregnant or breastfeeding; had a condition threatening life or compromising follow-up over the study period; had a comprehension deficit, defined as a score below 12/15 on the command-execution subtest of the Boston Diagnostic Aphasia Examination [3]; had fixed spasticity of the finger or wrist flexors, defined as a score of 4 on the modified Ashworth scale (mAS) [4], or had received botulinum toxin in the forearm or hand within the 12 weeks before the inclusion visit; had a history of degenerative neurological disease or craniectomy; had a skin lesion of the affected limb preventing electrical stimulation, or of the scalp preventing EEG placement; were taking part in another therapeutic study liable to affect recovery of the affected hand; had previously taken part in a brain-computer interface study.

##### S1.3 Recruitment

Patients were identified during routine follow-up in the rehabilitation departments of Pitié-Salpêtrière Hospital and the neurovascular department of the Fondation Adolphe de Rothschild Hospital, and were all recruited and included at the Paris Brain Institute on the day of the inclusion visit.

### S2 Initial assessment and BCI training

#### S2.1 Inclusion assessment

At the inclusion visit the eligibility criteria were verified and a fuller motor and cognitive assessment was administered, reported in Table 1. This assessment included the modified Rankin Scale (mRS) [5], the distal sensory subscale of the Fugl-Meyer assessment, the Montreal Cognitive Assessment (MoCA) [6], the Frontal Assessment Battery (FAB) [7], the Batterie d'Évaluation de la Négligence (BEN) for left spatial neglect [8], and the three subscales of the second French version of the Movement Imagery Questionnaire 3 (MIQ-3Sf) [9]. The weekly volume of usual physiotherapy was taken from the medical file.

#### S2.2 Calibration of the sensorimotor feedback

Before training, the sensorimotor feedback was calibrated to induce a standardised and functional hand opening. Stimulation was adjusted to activate preferentially the extensor digitorum communis, the extensor indicis proprius, and the abductor and extensor pollicis longus, while limiting wrist extension. Intensity was adapted individually according to the nociceptive threshold, pyramidal irritability, and the most natural and functional hand-opening pattern achievable for each patient. It was set at the lowest level producing a visible and complete opening of the hand without discomfort. The setting obtained at the start of a session was recorded and applied to every trial of that session.

As spasticity fluctuated with fatigue, stress and cognitive effort, the experimenter stretched the fingers to relieve it whenever the stimulation appeared visually less effective and sterelotyped.

### S3 Standardized guidance and interaction

All verbal exchanges between experimenter and patient before, during, and after runs are described in this section, organized chronologically within a session.

#### S3.1 Pre-run instructions

At the beginning of each session, the experimenter introduced the MI task using a standardized imagery script, designed to help participants learn, deepen, and refine their MI technique. Participants were encouraged to explore both kinesthetic and visual strategies and identify the sensory components that were most accessible and reproducible for them. A longer version was used for the first three sessions, followed by a shortened version thereafter.

##### S3.1.1 Long version

###### Grounding

Close your eyes and let yourself be guided by my voice. Take a moment to focus on the sensations throughout your body. Make yourself comfortable in the chair. Feel your back supported by the backrest and your arms resting comfortably on the armrests. Your feet are resting comfortably on the floor. Allow your body to become heavy and gradually relax, without any effort *[3 s]*.

#### Refocusing

Breathe in for as long as possible *[5 s]*, then breathe out gently *[5 s]*. Once again, breathe in and feel your abdomen expand *[5 s]*, then breathe out steadily *[5 s]*. Notice the cool air as it enters through your nose. Continue with this breathing pattern.

As you breathe out, your shoulders drop and the tension in your body disappears. With your next breath in, feel the cool air move down your throat and into your lungs. Then breathe out slowly. Feel your jaw relax, and your eyebrows soften *[2 s]*. With each breath, notice the gentle sensation of your clothes against your skin. With each breath, you become more and more relaxed *[2 s]*.

#### Connecting with the Healthy Hand and Motor Imagery Strategy

Keep your eyes closed and bring your attention to your *[right/left-healthy]* hand. Your hand is relaxed, and you can feel it resting against the cushion. Your fingers are relaxed, and you can feel a slight tension at your fingertips *[3 s]*. Now, try to feel these same sensations with your *[right/left-affected]* hand: the contact with the cushion, your fingers being relaxed, and the slight tension at your fingertips *[3 s]*.

We are now going to begin the motor imagery training. The goal is to create a precise mental image of your hand, opening to reach and grasp the cup on the table.

Open your eyes now. To help you imagine the movement, you will first perform it with your *[right/left-healthy]* hand. Open your *[right/left-healthy]* hand. Watch your fingers straighten and your thumb and fingers move apart. Keep your hand open and notice the activation across the back of your hand and the sensation in your skin. Keep this image in mind. Then, relax.

Now, imagine this movement with your *[right/left-healthy]* hand. You are relaxed, and none of the muscles in your *[right/left-healthy]* arm are moving. Imagine your hand slowly opening, just as you did, visualizing your fingers extending and feeling the tension as they move apart. Imagine keeping your hand open for as long as possible *[5 s]*. Then, relax.

We are now going to do the same exercise, but this time with your *[right/left-affected]* hand. To begin, we are going to send you a stimulation to help open your hand *[stimulation]*. Watch your fingers extend in front of you and move apart from one another. Keep this image in mind.

Now, imagine your hand slowly opening to reach and grasp for the cup, without actually moving. Visualize your thumb lifting and your fingers gradually spreading apart. Imagine all your fingers extending. Keep your hand open for as long as possible *[5 s]*. Then, relax and imagine your hand returning to its initial position.

Now, bring your attention to the sensations associated with the movement. Once again, we are going to send you a stimulation to help open your hand *[stimulation]*. Focus on the sensations in your hand as it opens. Feel your muscles contracting and the tension building in your hand.

Now, imagine these same sensations. In your forearm, feel your muscles contracting. Feel your tendons activate along the back of your hand. As the tension increases, imagine your thumb moving further away and your fingers lifting and extending. Keep your hand open for as long as possible *[5 s]* before relaxing.

##### Repetition

During the next stimulation, you can combine what you see with what you feel as your hand opens.

Keep your eyes open and imagine your hand opening once again. Try to include as many details as possible in your imagery *[5 s]*. Then, relax.

Are you ready? We are now going to begin the experiment.

##### S3.1.2 Short version

##### Grounding

Close your eyes and let yourself be guided by my voice. Take a moment to focus on the sensations throughout your body. Make yourself comfortable in the chair. Feel your back supported by the backrest and your arms resting comfortably on the armrests. Your feet are resting comfortably on the floor. Allow your body to become heavy and gradually relax, without any effort *[3 s]*.

##### Refocusing

Breathe in for as long as possible *[5 s]*, then breathe out gently *[5 s]*. Once again, breathe in and feel your abdomen expand *[5 s]*, then breathe out steadily *[5 s]*. Notice the cool air as it enters through your nose. Continue with this breathing pattern.

As you breathe out, your shoulders drop and the tension in your body disappears. With your next breath in, feel the cool air move down your throat and into your lungs. Then breathe out slowly. Feel your jaw relax, and your eyebrows soften *[2 s]*. With each breath, notice the gentle sensation of your clothes against your skin. With each breath, you become more and more relaxed *[2 s]*.

##### Connecting with the Hand and Motor Imagery Strategy

Keep your eyes closed and bring your attention to your *[right/left]* hand. Your hand is relaxed, and you can feel it resting against the cushion *[3 s]*. Your fingers are relaxed, and you can feel a slight tension at your fingertips *[3 s]*.

We are now going to begin motor imagery training. Open your eyes now. Focus on the cup placed on the table or, if it helps you, look at your *[right/left]* hand. The goal is to create a precise mental image of your hand, opening to reach and grasp the cup whilst keeping your arm relaxed and completely still.

Now imagine your hand slowly opening to grasp the cup. Visualize your thumb lifting and your fingers spreading from one another. All of your fingers are extended. Keep your hand open for as long as possible *[5 s]*. Then relax and imagine your hand returning to its initial position.

Now, focus on the sensations associated with the movement. Once again, imagine your hand opening. In your forearm, feel the muscles contracting and the tendons along the back of your hand activate. As the tension increases, your thumb moves further away, and your fingers lift and extend. Keep your hand open for as long as possible *[5 s]* before relaxing.

##### Repetition

Keep your eyes open throughout the exercise and imagine your hand opening once again. Try to include as many details as possible in your imagery *[5 s]*. Then relax.

Are you ready? We are now going to begin the experiment.

#### S3.2 Inter-run dialogue

Guidance and instructions were delivered by the same experimenter throughout the protocol. During BCI sessions, the experimenter adopted a standardized yet adaptive role in the development, monitoring, and refinement of participants' motor imagery (MI) strategies. The experimenter's involvement can be conceptualized across two stages: (1) bidirectional exchange and discussion following individual runs, and (2) block-level and end-of-session debriefing and retraining.

1. Run-by-run dialogue and adaptation

Following each experimental run, feedback was conceived as a bidirectional process, combining the participant's subjective experience with information derived from the BCI system. This provided complementary information about both how the participant performed the MI task and how their imagery was reflected in the recorded data.

First, the participant was invited to describe their experience of the MI task using open-ended questions. Discussions addressed how the participant experienced the task, which MI strategy they used including the relative contribution of visual and kinesthetic components and whether their strategy had changed.

Second, the experimenter provided feedback based on information obtained during the run. These included online linear discriminant analysis (LDA) classification and confidence scores, changes in classifier features, and the recorded EEG signal, together with video and EMG recordings used to identify overt hand movements, muscle contractions, or other potential sources of artifact. Offline processing of the preceding run was additionally used to examine $R^2$ maps and spectral profiles during rest and MI conditions, allowing changes in the spatial distribution of activity at the electrode level and in frequency-domain characteristics to be examined over time.

The experimenter then integrated these sources of information to test and update hypotheses regarding the participant's MI strategy. For example, a change in the resting-state signal could prompt discussion about whether the participant had been reflecting on the task between runs, while a change in the spatial or spectral characteristics of the EEG signal could prompt exploration of whether the participant had modified their MI strategy.

This process allowed the experimenter to determine whether the current MI strategy should be maintained, modified, or further developed. When classification performance was poor or unstable, the MI strategy could be reconsidered or simplified. When performance was good and stable, the strategy could be progressed by encouraging greater vividness. Thus, feedback was not limited to informing the participant of BCI performance, but was used as part of an iterative process of hypothesis generation, testing, and adaptation.

Participant state was also monitored. Where increased effort, stress, or fatigue appeared to be associated with increased muscle tension or spasticity, stretching, or rest periods were introduced. A camera was additionally used to monitor the FESIA device and the quality of hand opening, allowing stimulation or positioning to be adjusted as necessary.

2. Block-level debriefing, retraining, and session conclusion

In addition to the brief discussions conducted after individual runs, more extensive debriefing was performed at the end of blocks, during periods in which the BCI classifier was being retrained.

During these periods, the experimenter and participant could develop updated hypotheses regarding the most effective MI strategy, informed by the convergence between the participant's reported experience and observed changes in the EEG signal. Where appropriate, FESIA stimulation was used again to help anchor or reinforce the sensory experience associated with the intended movement.

A final debriefing was conducted at the end of each session to review the participant's overall experience, identify difficulties, their perceived sense of control and provide an opportunity for questions and feedback.

[6] Nasreddine ZS, Phillips NA, Bédirian V, Charbonneau S, Whitehead V, Collin I, et al. The Montreal Cognitive Assessment, MoCA: A Brief Screening Tool For

Mild Cognitive Impairment. Journal of the American Geriatrics Society. 2005 Apr;53(4):695–699. https://doi.org/10.1111/j.1532-5415.2005.53221.x.

[7] Dubois B, Slachevsky A, Litvan I, Pillon B. The FAB: A frontal assessment battery at bedside. Neurology. 2000 Dec;55(11):1621–1626. https://doi.org/10.1212/WNL.55.11.1621.

[8] Azouvi P. Sensitivity of clinical and behavioural tests of spatial neglect after right hemisphere stroke. Journal of Neurology, Neurosurgery & Psychiatry. 2002 Aug;73(2):160–166. https://doi.org/10.1136/jnnp.73.2.160.

[9] Robin N, Coudevylle GR, Dominique L, Rulleau T, Champagne R, Guillot A, et al. Translation and validation of the movement imagery questionnaire-3 second French version. Journal of Bodywork and Movement Therapies. 2021 Oct;28:540–546. https://doi.org/10.1016/j.jbmt.2021.09.004.
