## Supplementary material for "Single-Case Design for Network-based BCI Rehabilitation: Individual Learning Trajectories and Effects of Motor Imagery Timing in Stroke Patients": Supp Tables

Table S1: Clusters of longitudinal change and of motor imagery time, by patient, effect and estimator.

| Patient | Effect | Est. | Electrodes (by peak) | Band | Bins | Slope | $p$ | Min | Max | Mean | $g$ |
|---|---|---|---|---|---|---|---|---|---|---|---|
| P1 | Session | $\phi$ | C5 FC5 | 12-18 | 6 | +4.537 | .019 | +4.036 | +5.540 | +4.788 | 1.86 |
| | | $\phi$ | C4 CP4 C2 CP2 | 28-30 | 6 | +9.231 | <.001 | +5.875 | +11.451 | +8.562 | 3.37 |
| | | $s$ | CP5 C3 | 8-12 | 4 | +0.729 | .028 | +0.525 | +0.933 | +0.729 | 1.60 |
| | | $s$ | FC6 FC4 C5 C2 FC5 CP5 CP2 C3 C4 CPz CP6 | 22-30 | 22 | +1.934 | .003 | +0.586 | +3.048 | +1.766 | 1.08 |
| | MIT | $\phi$ | *no cluster* | - | - | - | - | - | - | - | - |
| | | $s$ | *no cluster* | - | - | - | - | - | - | - | - |
| P2 | Session | $\phi$ | CPz CP1 CP2 FC1 C3 Cz C4 C2 C1 CP4 CP3 CP5 FC3 C5 | 8-24 | 43 | +4.505 | .009 | +1.948 | +12.474 | +4.762 | 0.85 |
| | | $s$ | FC1 CPz FC5 FC6 FCz CP4 C4 | 8-12 | 12 | +3.217 | .032 | +2.028 | +4.406 | +3.271 | 1.36 |
| | | $s$ | CP3 CPz CP1 FC6 C1 C5 C3 FC4 CP5 FC5 FC3 | 16-30 | 43 | +2.341 | .014 | +0.900 | +3.588 | +2.222 | 0.77 |
| | MIT | $\phi$ | *no cluster* | - | - | - | - | - | - | - | - |
| | | $s$ | *no cluster* | - | - | - | - | - | - | - | - |
| P3 | Session | $\phi$ | FCz Cz CP2 C2 CPz FC2 C4 CP4 | 12-30 | 35 | +11.323 | <.001 | +4.117 | +17.663 | +10.634 | 1.44 |
| | | $s$ | FCz C2 CPz Cz FC2 C1 CP1 CP2 FC3 FC5 FC1 | 16-26 | 34 | +10.044 | .001 | +6.416 | +13.901 | +9.526 | 1.89 |
| | MIT | $\phi$ | *no cluster* | - | - | - | - | - | - | - | - |
| | | $s$ | *no cluster* | - | - | - | - | - | - | - | - |
| P4 | Session | $\phi$ | C2 Cz C1 CP3 FCz CP5 C3 CP1 CP2 FC1 C5 CPz C4 CP4 C6 FC2 FC3 CP6 FC6 FC4 | 8-30 | 165 | +11.354 | <.001 | +6.920 | +15.487 | +10.969 | 0.79 |
| | | $s$ | C1 Cz FCz C3 C2 CP2 FC1 CPz FC5 FC3 CP1 CP5 CP4 | 20-26 | 25 | +2.185 | .002 | +0.803 | +3.945 | +1.841 | 1.32 |
| | MIT | $\phi$ | C3 FCz FC1 FC3 FC5 | 12-26 | 20 | −52.009 | .013 | −61.273 | −48.202 | −54.024 | −1.15 |
| | | $s$ | C1 CPz Cz | 8-10 | 4 | −8.539 | .026 | −11.153 | −5.742 | −7.668 | −1.33 |
| P5 | Session | $\phi$ | FCz FC1 FC2 C3 FC3 CP5 | 8-12 | 11 | +9.789 | .020 | +8.423 | +11.596 | +9.512 | 1.80 |
| | | $s$ | *no cluster* | - | - | - | - | - | - | - | - |
| | MIT | $\phi$ | *no cluster* | - | - | - | - | - | - | - | - |
| | | $s$ | *no cluster* | - | - | - | - | - | - | - | - |
